# MASCOT-DS improves transmission dynamics inference by integrating multiple epidemiological data streams with phylodynamic inference

**DOI:** 10.64898/2026.08.21.26361056

**Authors:** Paula H. Weidemüller, Luis R. Esquivel Gomez, Isabel Rodriguez-Barraquer, Nicola F. Müller

## Abstract

Tracking how an infectious disease spreads in time and space relies on several distinct sources of surveillance data, reported case counts, viral concentrations in wastewater, seroprevalence surveys, and pathogen genomic sequences, each of which is imperfect and captures only part of the underlying transmission process. These data streams are typically analyzed separately or with highly parameterized, disease-specific models, making it difficult to combine their complementary strengths. Here we present MASCOT-DataStreams (MASCOT-DS), a BEAST2 software package that extends the structured coalescent model MASCOT to jointly infer prevalence over time and transmission rates between locations from any combination of case counts, wastewater concentrations, seroprevalence surveys, and pathogen phylogenies. Using simulated outbreaks in structured populations, we show that MASCOT-DS accurately recovers true prevalence trajectories and between-location migration rates. We then apply MASCOT-DS to genomic, case count, wastewater, and seroprevalence data from the SARS-CoV-2 Epsilon wave (winter 2020-21) in three San Francisco Bay Area counties, reconstructing county-level prevalence dynamics and quantifying transmission within and into the region. By systematically removing individual data streams, we find that genomic data are uniquely required to estimate transmission between locations, while seroprevalence data are essential for anchoring the overall magnitude of an outbreak; case counts and wastewater concentrations play largely interchangeable roles in capturing outbreak shape. These results demonstrate that integrating complementary epidemiological data streams substantially increases the certainty of transmission dynamics estimates compared to relying on any single data stream, and provides a framework for evaluating the added value of different surveillance strategies.

**Significance:** Understanding how infectious diseases spread between communities is crucial for public health responses, but no single surveillance method such as case reporting, wastewater monitoring, serosurveillance, or genome sequencing is able to fully inform all aspects of pathogen transmission dynamics. We developed MASCOT-DS, a phylodynamic model that jointly infers transmission dynamics from data streams, applied here to the SARS-CoV-2 Epsilon wave in the San Francisco Bay Area. Combining data streams produced more reliable estimates than any single source, and each data type provided distinct, complementary information. This work offers a general framework for robust pathogen transmission dynamics inference and helps public health agencies evaluate and optimize how their surveillance data informs transmission dynamics reconstruction.

## Introduction

Traditionally, transmission dynamics of infectious diseases have been inferred from time series of reported case counts and, where available, seroprevalence studies using epidemiological models. Over the past decades, increasing capacity and decreasing costs of sequencing have made routine pathogen sequencing feasible, enabling the analysis of genomic data with phylodynamic methods that reconstruct pathogen spread through space and time [1, 2, 3]. More recently, wastewater surveillance has become routine in many regions, where pathogen concentrations are regularly measured and often made publicly available in near-real time through efforts such as WastewaterSCAN [4, 5, 6]. Even with significant growth in data availability associated with expanded surveillance infrastructure and reporting systems during the SARS-CoV-2 pandemic, methods for transmission dynamics reconstruction are still typically developed in isolation for individual data streams or just a subset of them, often requiring the use of highly parameterized mechanistic models [7, 8, 9, 10, 11, 12, 13, 14, 15, 16, 17, 18, 19]. Because each data stream captures different aspects of the underlying transmission dynamics, models integrating multiple streams could yield more precise estimates of spatiotemporal disease prevalence and between-location transmission rates. We subsequently refer to how prevalence changes over time in a given location as the “prevalence trajectory”.

In this study, we considered three epidemiological data streams. Case counts, which capture a fraction of symptomatic infections at different timepoints, provide information about the shape and trends of the prevalence trajectory. However, due to variable symptomatic fractions, under-ascertainment, biases in testing over time and between locations and reporting delays, accurately quantifying the actual magnitude of prevalence over time is difficult [20]. Wastewater pathogen concentrations, which measures the concentration of pathogen DNA in sewage, can provide information on the temporal dynamics of an outbreak that are not subject to the biases of passive surveillance systems such as variable and unreliable case ascertainment and reporting delays [21, 22, 23, 24, 17]. However, calibrating viral concentrations in wastewater to actual prevalence is difficult based on wastewater surveillance alone, and measurements of viral concentrations in sewage can be very noisy in part due to high variance in shedding between individuals and along the schedding period, dilution effects and sampling methodology [4, 25, 26, 27, 28, 22, 29, 30, 17]. Seroprevalence observations, which represent the number of people with antibodies against a specific pathogen, provide information on the overall magnitude of an outbreak, such as the cumulative proportion of a population that has been infected by a specific point in time [31, 32, 33, 34]. Seroprevalence estimates can be obtained through population-wide seroprevalence studies, but these are rarely and infrequently performed, limiting the use of this data stream in resolving prevalence trajectories over multiple timepoints. They can also be obtained by testing subsets of residual blood samples obtained from blood donors or patients, though these may be less representative of the general population.

Genomic pathogen sequences, which capture the evolutionary relationships between cases, provide an excellent data source to study spatial transmission dynamics using phylodynamic methods, as they enable us to link cases across time and space using the mutational relatedness between sampled sequences to construct phylogenetic trees. Unlike any of the other data streams above, these trees allow estimation of “migration rates”, that quantify pathogen transmission between locations. However, in practice, only a subset of infections is sequenced, and sequencing effort can vary over time and between locations. Phylodynamic analyses become increasingly computationally demanding as the number of sequences grows or as population structure becomes more complex, e.g. increasing the number of locations of interest or temporal changes in population size, limiting the size and scope of datasets that can be analyzed [35, 36, 37]. Furthermore, it is difficult to infer the exact shape and magnitude of prevalence trajectories from phylogenetic trees, especially in complex population structures and without additional data [7, 8, 38, 39, 3].

In this work, we describe MASCOT-DataStreams (MASCOT-DS), a BEAST2 [40] package that extends the structured coalescent model MASCOT [41, 42] to optionally incorporate case counts, wastewater viral concentrations, and serological data as independent likelihood terms to jointly infer prevalence trajectories and migration rates in structured populations, defined as populations partitioned into distinct subgroups such as different locations. We first validated MASCOT-DS using simulated outbreaks, then applied it to genomic, case count, wastewater, and seroprevalence data from the SARS-CoV-2 Epsilon wave across three San Francisco Bay Area counties in winter 2020-21, inferring county-level prevalence trajectories and transmission dynamics both between counties and from outside the region. Last, we examined the contribution of each data stream to the transmission dynamics estimates. By integrating multiple epidemiological data streams within a unified framework, MASCOT-DS leverages the complementary strengths of each data source to improve inference beyond what is possible using any single data stream.

## Results

### MASCOT-DS integrates epidemiological data streams into structured phylodynamic inference

We model prevalence trajectories and pathogen migration rates in structured populations by incorporating various epidemiological data streams into the Bayesian phylodynamic model MASCOT-DS. In order to jointly infer prevalence trajectories *I*(*t*) and pathogen migration rates *m*, we model each data type with an independent, data stream-specific likelihood term *P* (*D*|*I*(*t*), …). The term denotes the probability of observing the data under the model given parameters such as *I*(*t*), which are estimated through Markov Chain Monte Carlo (MCMC) sampling within the BEAST2 software. Each likelihood function is defined by a different probability distribution parametrized with certain parameters, such as scaling factors *k*, that are estimated alongside the parameters of interest, *I*(*t*) and *m*. We model the phylogeny, which refers to the phylogenetic trees, with the MASCOT structured coalescent likelihood, case counts (and any discrete, non-negative data) with a negative binomial, wastewater pathogen concentrations (and any continuous, strictly-positive data) with a log-normal and seroprevalence observations with a binomial likelihood function. Figure 1A and Table 1 provide an overview over the different likelihoods and parameters used in the model.

**Table 1:**
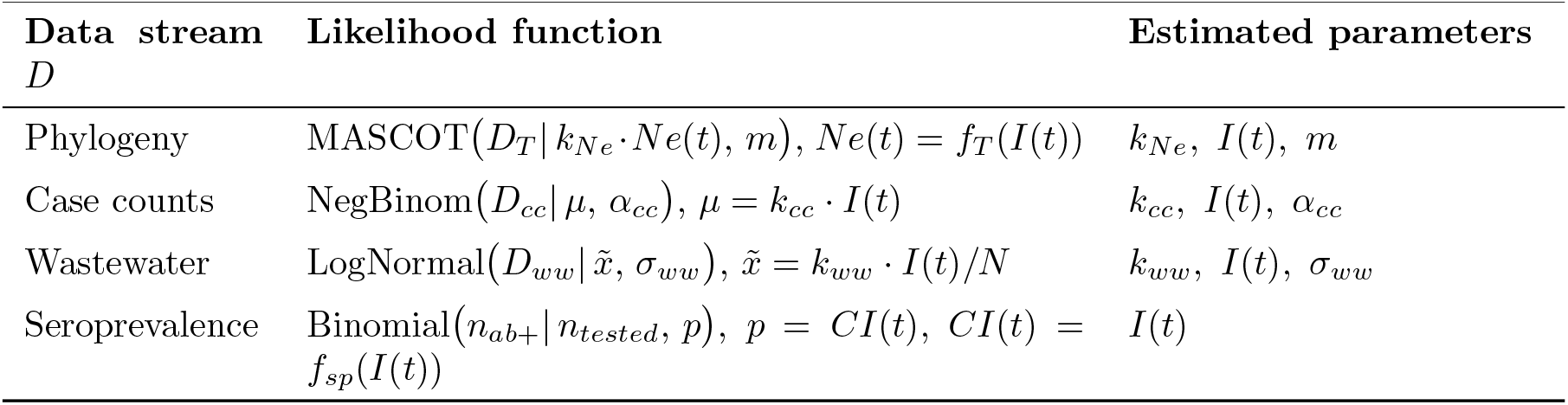
Data streams *D*, likelihoods, and estimated parameters. Input data are: the phylogeny *D*_*T*_, case counts *D*_*cc*_, wastewater pathogen concentrations *D*_*ww*_, seroprevalence observations *D*_*sp*_ with *n*_*tested*_ being the number of people tested and *n*_*ab*+_ being the number of people with antibodies against a pathogen. Parameters are: *µ* is the mean, 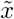 is the median, *α*_*cc*_ is dispersion, *σ*_*ww*_ is the standard deviation, *k* are scaling factors, *I*(*t*) are location-specific prevalence trajectories and *m* are the migration rates between locations. *CI* is the cumulative incidence derived from *I* (*f*_*T*_ (*I*)) and *Ne* is the effective population size derived from *I* (*f*_*sp*_(*I*)).

**Figure 1:**
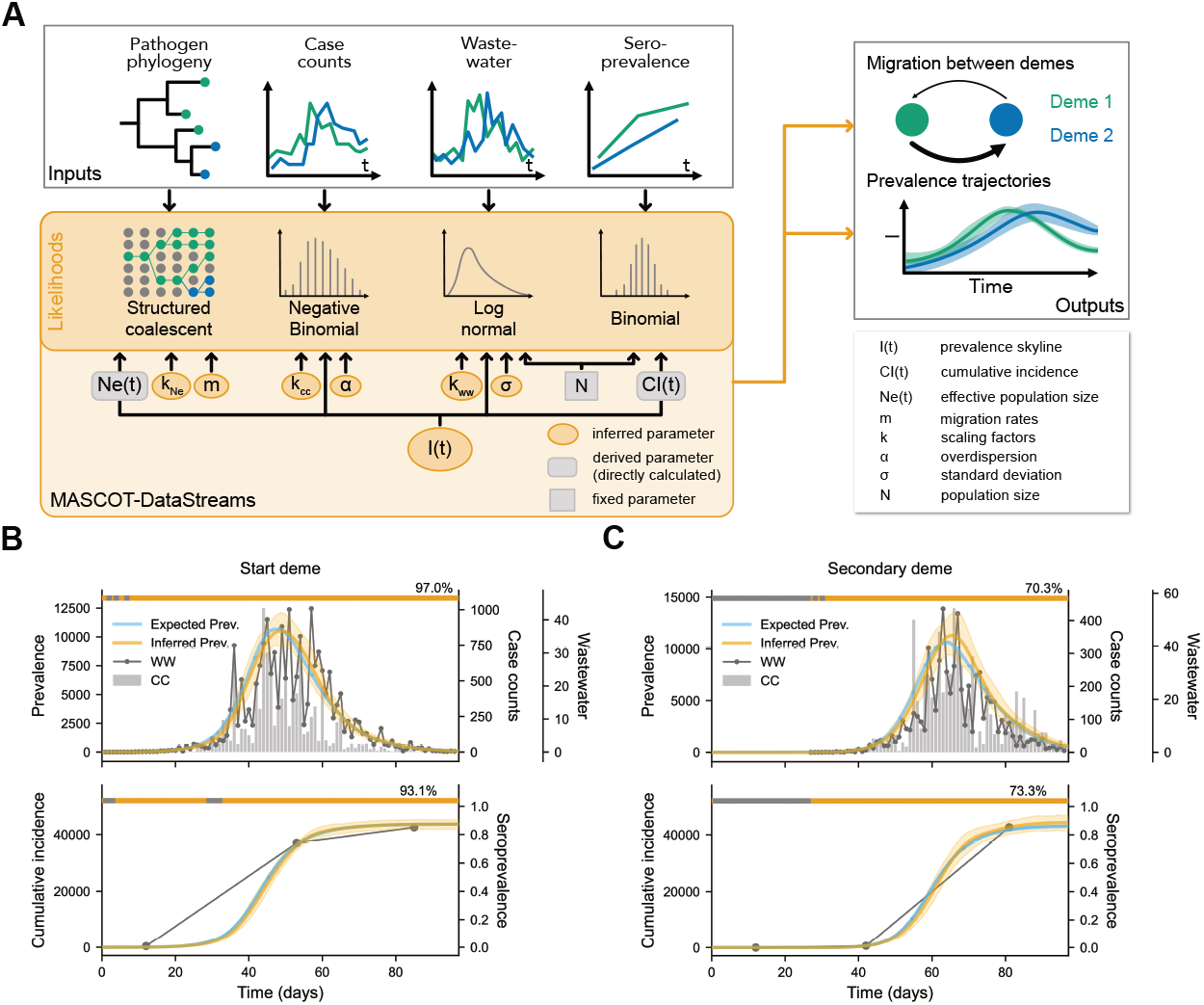
Overview of the MASCOT-DataStreams (MASCOT-DS) model implemented as a BEAST2 package. A) Schematic of the MASCOT-DS data inputs *D*, likelihoods *P* (*D*|…) and estimated parameters. Also see Table 1. The phylogenetic tree (phylogeny) is a direct input into MASCOT-DS, but using other functions available in the wider BEAST2 ecosystem, the user can also input a sequence alignment from which the phylogeny is estimated. B&C) Example of MASCOT-DS estimates on a simulated SIR outbreak starting in the ‘start deme’ and spreading to the ‘secondary deme’ showing inferred prevalence and cumulative incidence (median of posterior as orange, solid lines; 95% highest posterior density (HPD) interval as orange shaded area). The true trajectories are shown as blue solid lines. The simulated wastewater concentrations and seroprevalence measurements are shown as dots and case counts as bars. In addition, MASCOT-DS was provided with the simulated phylogenetic tree. The bar on top of each plot indicates the percentage of timepoints in which the true prevalence/cumulative incidence value was contained in the 95% HPD interval. Where true prevalence was 0 no wastewater concentrations were simulated.

To validate the ability of MASCOT-DS to recover structured transmission dynamics, we performed a simulation study. In total, we ran 100 outbreak simulations in two synthetic demes using a SIR compartmental model, with varying transmission rates and recovery rates to cover a range of outbreak scenarios. Per simulation this produced one ground truth phylogenetic tree and prevalence trajectory per deme. We define demes as discrete subpopulations, within which transmission dynamics are assumed homogeneous (e.g. in a specific location), coupled by migration rates representing transmission between demes. From the simulated prevalence trajectories, we then generated synthetic case counts, wastewater concentrations and seroprevalence observations by sampling the data stream parameters, such as scaling factors, from fixed prior distributions and using the data stream-specific probability distributions to generate each data stream (Methods, Table 1). One thing to note, is that in MASCOT-DS, we currently assume no relevant detection limit of viral particles in wastewater and don’t support using concentrations of 0 as input, thus, in the simulation we did not provide wastewater measurements at timepoints where the true prevalence was 0. In reality, a measurement of 0 still carries information, as it can imply true absence of viral particles or concentrations below the assay detection limit [4]. Future improvements could extend both the case count and wastewater likelihood terms with a zero-inflated model, to more accurately model wastewater data but also case counts in scenarios where pathogen transmission can truly die out in a location (resulting in long stretches of 0 case counts).

We then provided MASCOT-DS with the simulated data streams including the fixed, corresponding phylogenetic tree and ran MCMC inference to obtain posterior distributions of prevalence trajectories, migration rates and data stream-specific likelihood parameters for each simulation. We did not estimate the tree in BEAST2 here, since our focus was on evaluating how different data streams affect transmission parameter estimates from MASCOT-DS and our simulations provided the ground truth tree. Trees can also be estimated from pathogen sequences using other functions in the BEAST2 ecosystem, then coupled with MASCOT-DS.

Figure 1B&C shows the 95% highest posterior density (HPD) interval of inferred prevalence and and cumulative incidence trajectories for an individual simulation alongside the input epidemiological data streams, both for the start deme (where the outbreak originated) and the secondary deme (where the outbreak spread to). Across simulations, MASCOT-DS was largely able to correctly reconstruct the prevalence values for the starting (Fig. 2A) and secondary deme (Fig. S2A) achieving a minimum of 50% and maximum of 99% coverage of the true prevalence values in the −95% HPD interval (Fig. 2B& S2B). Coverage was considerably better during the active outbreak period at high prevalence values compared to the start of the outbreak with no strong bias observed across the outbreak trajectory (Fig. 2C& S2C). MASCOT-DS recovered the true parameters of case count and wastewater likelihoods (Fig. 2D), with a coverage of 84% − 95.5% depending on parameter (Fig. 2E). The 84% coverage of the wastewater standard deviation *σ*_*ww*_ was lower than expected and slightly overestimated by MASCOT-DS (Fig. 2F). A posterior predictive check showed the predictive wastewater concentration distribution was well-calibrated, achieving near-nominal (≈95%) coverage of the observations (Fig. S3A), indicating that the model adequately captured the properties of the wastewater data. We consider the slight overestimation of *σ*_*ww*_ to be due to the inability of the prevalence spline approximation to fit the stochastic and discrete prevalence values at the start of an outbreak, which led to an inflated estimate of the noisiness of the wastewater observations at low prevalence (Fig. S3B&C).

**Figure 2:**
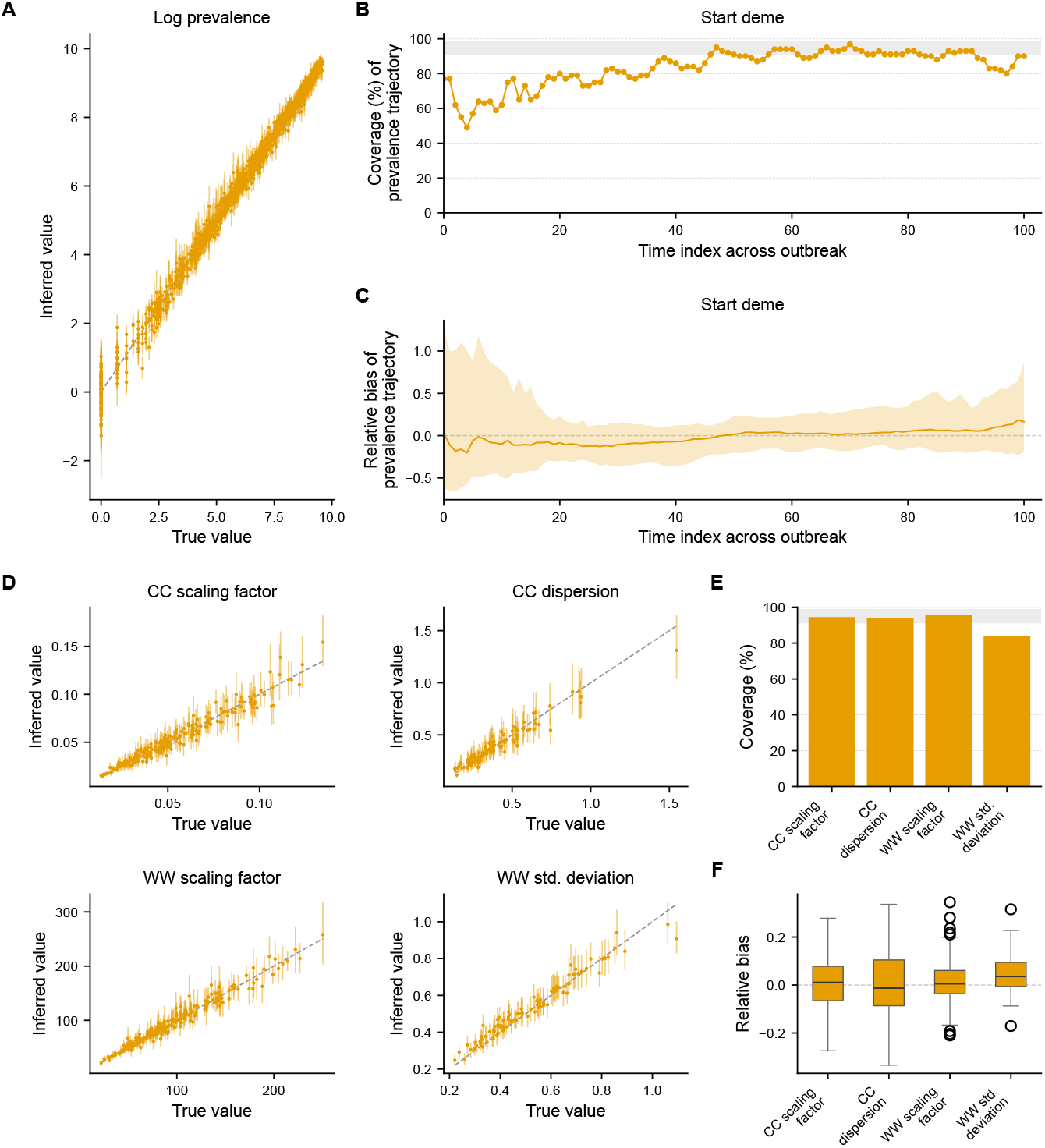
MASCOT-DS validation using 100 two-deme SIR simulations. A) Scatter plot of the inferred log prevalence values against the true log-transformed prevalence values for the starting deme across 100 simulations. Each dot is the median of the posterior of one of 11 knot points of the prevalence trajectory of a given simulation, whiskers span the 95% HPD interval. B) Coverage of true prevalence values in the 95% HPD interval of the estimated prevalence trajectory across all simulations. Grey band indicates desired 91-99% coverage. C) Relative bias of prevalence estimates relative to the true prevalence values. For each simulation, the true value was subtracted from the median of the posterior distribution and divided by the true value. The line shows the median bias across simulations, shaded area the 2.5 and 97.5 percentiles. D) Scatter plots of the inferred case count (CC) and wastewater (WW) likelihood parameters across 100 simulations. E) Coverage of the true CC and WW likelihood parameters across 100 simulations. Grey band indicates 91-99% coverage. F) Relative bias of CC and WW likelihood parameter estimates relative to the true parameters. The box plots summarize the relative bias across 100 simulations (box = IQR, line = median, whiskers = 1.5*×*IQR, points = outliers).

MASCOT-DS correctly estimated migration rates in the direction from the starting deme to the secondary deme with no bias, while it was not able to discern the magnitude of migration from the secondary deme to the starting deme (Fig. S2D&F) and thus simply sampling from the prior, due to very few migration events from the secondary to the start demes in the simulated trees. Still, the true values of both migration directions were contained in the 95% HPD in 93-94% of the simulations (Fig. S2E), indicating a well-calibrated model.

### MASCOT-DS reconstructs spatiotemporal transmission dynamics of SARS-CoV-2 Epsilon wave in the San Francisco Bay Area

To test the ability of MASCOT-DS to reconstruct pathogen transmission dynamics using real-world data, we applied MASCOT-DS to data from the SARS-CoV-2 wave (winter 2020-21) in the San Francisco Bay Area. We picked this wave, since it was largely driven by the Epsilon variant (B.1.427&B.1.429 lineages), which was responsible for much of the SARS-CoV-2 cases in California [43, 44, 45], surging from 2.1% in the week of September 28, 2020 to its peak of 60.5% during the week of February 15, 2021 [46]. We chose the three Bay Area counties, Sacramento, San Francisco and Santa Clara (which includes San Jose as the largest city in the Bay Area), because they are geographically spread but interconnected. All had consistent genomic, case and wastewater surveillance which presented an ideal test case scenario for applying MASCOT-DS to real world data. In addition, most people in California had not been exposed to SARS-CoV-2 before the Epsilon wave (seroprevalence of 4.5% on October 13, 2020, Fig. 3B right), which meant we didn’t have to account for substantial prior infections. Despite the winter 2020-21 SARS-CoV-2 wave being driven by the Epsilon variant, case counts, wastewater concentrations and seroprevalence likely captured the spread of all circulating variants, while we restricted the phylogeny to only Epsilon sequences for computational efficiency. Since we expect the underlying population structure of the viral spread to be the similar enough between relevant variants, we assume that the Epsilon phylogeny still accurately reflected the overall SARS-CoV-2 transmission dynamics during the winter 2020-21 wave in California.

**Figure 3:**
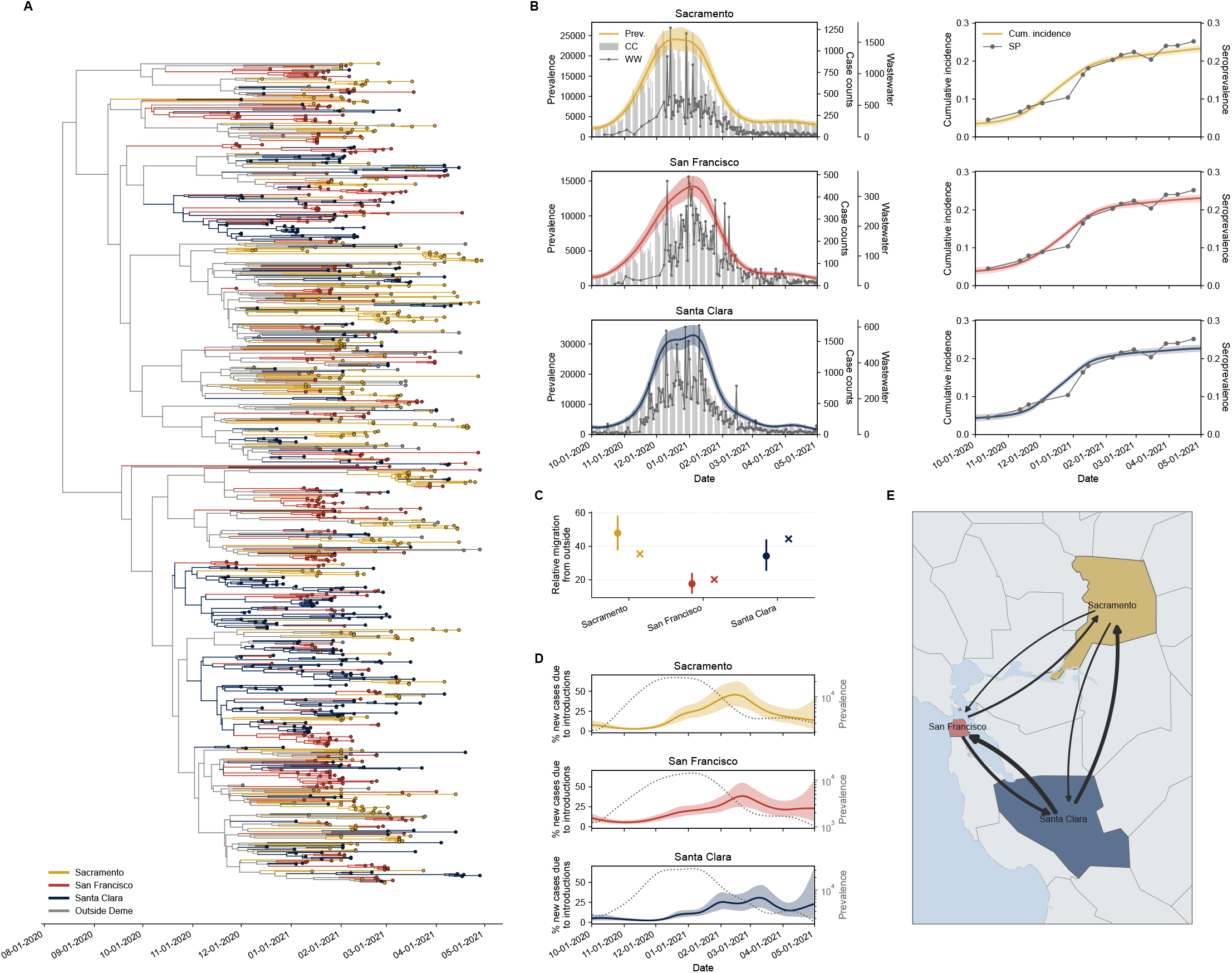
Reconstruction of transmission dynamics of the SARS-CoV-2 winter 2020-21 wave in the San Francisco Bay Area. A) Phylogenetic tree inferred by MASCOT-DS based on Epsilon variant sequences, case counts, wastewater concentrations and seroprevalence observations. The posterior distribution of trees was summarized into the maximum clade credibility tree keeping its node heights, lineages are colored by the location with the highest posterior probability of their corresponding child node. B) Inferred prevalence (prev.) and cumulative (cum.) incidence trajectories for the three Bay Area counties. Colored, solid lines show the median of the posterior distribution, the shaded area shows the 95% HPD interval. Case counts (CC) are shown as gray bars, wastewater concentrations (WW) and seroprevalence observations (SP) are shown as gray dots connected by lines. C) Inferred migration rates from the outside deme into the counties relative to each other; dots are the median of the posterior distribution, whiskers span the 95% HDP interval. The “X” indicates relative population sizes of each county for comparison. D) Percentages of new cases due to introductions from the outside deme over time. Dotted lines show the prevalence trajectory of a county, same as shown in B). E) Migration rates between local demes, width of arrows is scaled by the median inferred migration rate (Fig. S4).

We constructed a 999 sequence multiple sequence alignment with 310 randomly selected Epsilon sequences of each county and 70 randomly selected sequences across the US, which served as an evolutionary background from which lineages could migrate into our three counties of interest, we call this background the “outside deme”. All counties reported daily case counts and near-daily wastewater concentrations. In the absence of county-specific seroprevalence surveys, we instead used the California wide Commercial Laboratory Seroprevalence Survey as stand-in surveys for each deme. This was done to provide information on the overall magnitude of the wave, since we assumed that the level and timing of exposure to SARS-CoV-2 was similar across counties. This decision comes with two caveats. Firstly, using the same data for each county as independent observations likely overestimated the certainty of the parameter estimations. Secondly, this forced MASCOT-DS to estimate a similar cumulative incidence over the course of the wave for each county.

Figure 3A shows the maximum clade credibility tree with branch colors indicating the maximum posterior location estimate. The Epsilon variant was introduced into the different San Francisco Bay Area counties multiple times from outside and showed also frequent exchange between the counties instead of forming distinct county-specific clades (Fig. 3A). MASCOT-DS estimated that the prevalence peaked in late December 2020 to early January 2021 in all three counties (Sacramento: 24,104 (95% HPD interval: 21,683–26,629) infections on December 17, San Francisco: 14,224 (12,882–15,656) infections on January 4, Santa Clara: 32,905 (29,857–36,178) infections on January 4), consistent with the high case counts and wastewater viral concentrations in this period (Fig. 3B left). Since all three counties shared the same seroprevalence observations, the cumulative incidence at the end of the time period of interest (2021-04-30) was estimated to be 23.3% (95% HPD interval: 22.4-24.1%) for Sacramento, 23.1% (22.2-23.9%) for San Francisco and 22.7% (21.9-23.5%) for Santa Clara (Fig. 3B right).

Migration rates from the outside into the counties were proportional to each county’s population size (Fig. 3C), implying an overall similar proportion of cases due to introductions in each county. The Epsilon wave in each county was mainly driven by local transmission, with introductions from the other counties and the outside playing a larger role only during the downturn of the outbreak (Fig. 3D).

Sacramento showed lower migration rates to the other two counties, while the highest migration rate was estimated from Santa Clara to San Francisco (Fig. 3E, Fig. S4). However, HPD intervals overlapped considerably across migration routes (Fig. S4), still Santa Clara was estimated source of highest transmission to the other two counties. Since seroprevalence observations are important in setting the overall magnitude of the prevalence, we performed a sensitivity analysis, where we artificially doubled or halved the values of the seroprevalence observations. As expected this increased and decreased the overall estimated magnitude of the prevalence trajectory, respectively (Fig. S5A). With increasing values of seroprevalence the estimated number of migration events from outside into the counties decreased slightly, and more migration events between the counties were inferred (Fig. S5B,C). Importantly, the estimated relative migration from outside into each county was robust to the scale of seroprevalence values (Fig. S5D). The temporal dynamics of the percent of new cases due to introductions was equally robust, with only slight variation in the overall estimated magnitude (Fig. S5E).

### Complementary data streams each contribute distinct information to phylodynamic inference

Having a unified model allowed us to test the contribution of each data stream to parameter inference by performing a leave-one-out analysis. To this end, we repeated the simulation study and SARS-CoV-2 analysis in five different configurations of input data. First, we only provided the phylogeny (“Phylogeny only”), which is closest to standard phylodynamic methods. For the other four configurations we each omitted one source of information (“No case counts”, “No wastewater”, “No seroprevalence”, “No phylogeny”) while retaining the other remaining data streams. In the case of simulated data we compared these configurations + the full data streams model (“All DS”) to the ground truth, in the case of the SARS-CoV-2 analysis we compared the resulting parameter inferences to the “All DS” version.

Focusing on the simulations, removing case counts or the phylogeny did not change prevalence trajectory inference. Removing wastewater concentrations led to an underestimation of prevalence associated with increased uncertainty at the beginning of the outbreak (Fig. S6A). When seroprevalence observations were removed, prevalence estimation was more uncertain and biased towards larger values (Fig. S6A), which was associated with more uncertain and systematically underestimated scaling factors of the case counts and wastewater concentration likelihoods (Fig. S8A,C). When MASCOT-DS was only given the phylogenetic tree as input, prevalence estimation was highly uncertain and did not capture the downturn of the outbreak (Fig. S7A). The same patterns were visible in the secondary deme (Fig. S7B). Migration rate inference was slightly biased upwards with larger uncertainty when seroprevalence observations were removed. As expected, when the phylogenetic tree was removed (Fig. S6B and Fig. S7C), migration rates were sampled from the prior distribution since only the phylogenetic tree can provide information of transmission between demes through the ancestral relationships between genomic samples. However, adding the epidemiological data streams to the phylogeny as input reduced uncertainty in the exact migration rate estimates (compare “All DS” vs. “Phylogeny only”, Fig. S6B and Fig. S7C), highlighting that more precise estimates of prevalence within demes also improve transmission estimates between demes. In terms of computational run time, adding more data increased the number of samples and burn-in time of the posterior chain compared to providing just the phylogeny (“Phylogeny only”, Fig. S9A,B). Computing the coalescent likelihood took up most of the burn-in time as could be seen by a reduction in the wall clock time until stationarity when just the epidemiological parameters were provided (“no phylogeny”) (Fig. S9B). It also took more time to reach convergence (effective sample size=200) of the posterior when all data was given compared to just providing the epidemiological parameters or only the phylogeny (Fig. S9C). However, this small increase in run time was offset by improved certainty of parameter estimates as discussed earlier and also seen in an increase in the certainty of the posterior (Fig. S9D).

Using the example of the SARS-CoV-2 winter 2020-21 wave in the San Francisco Bay Area, we saw similar patterns (Fig. 4). The phylogeny alone produced more uncertain estimates of migration events and prevalence trajectories and did not capture the decrease in cases (Fig. 4A,C and Fig. S10). Removing seroprevalence led to increased uncertainty of prevalence, total migration events and scaling factor estimates (Fig. 4B,C, S10, and S12). Overall less local migration events and more migration from the outside deme were estimated (Fig. 4C, S10C, and S11). Without seroprevalence data there was no information about the absolute magnitude of the outbreak which leads to non-identifiability between the case count, wastewater and effective population size scaling factors together and the overall scale of the prevalence trajectory. Thus, we also tested the results when fixing the case count or effective population size scaling to the posterior median of the “All DS” model. Estimates resembled the “All DS” model closest when fixing the case count scaling (Fig. 4B), in contrast fixing effective population size scaling resulted in less certain and lower prevalence estimates though less pronounced than in the complete “No seroprevalence” variant, total migration events locally and from outside were most uncertain for this version (Fig. 4C and S10C). Removing seroprevalence observations was also associated with a lower posterior estimate of the case count dispersion parameter (Fig. S12). Without seroprevalence data to constrain the prevalence trajectory, the model likely fit the shape of the prevalence trajectory closer to the case count observations, requiring less dispersion to capture the case count dynamics. Instead, the large 95% HPD interval of the case count and wastewater scaling factors and throughout the prevalence trajectory showed great uncertainty in the overall magnitude of prevalence rather than its shape.

**Figure 4:**
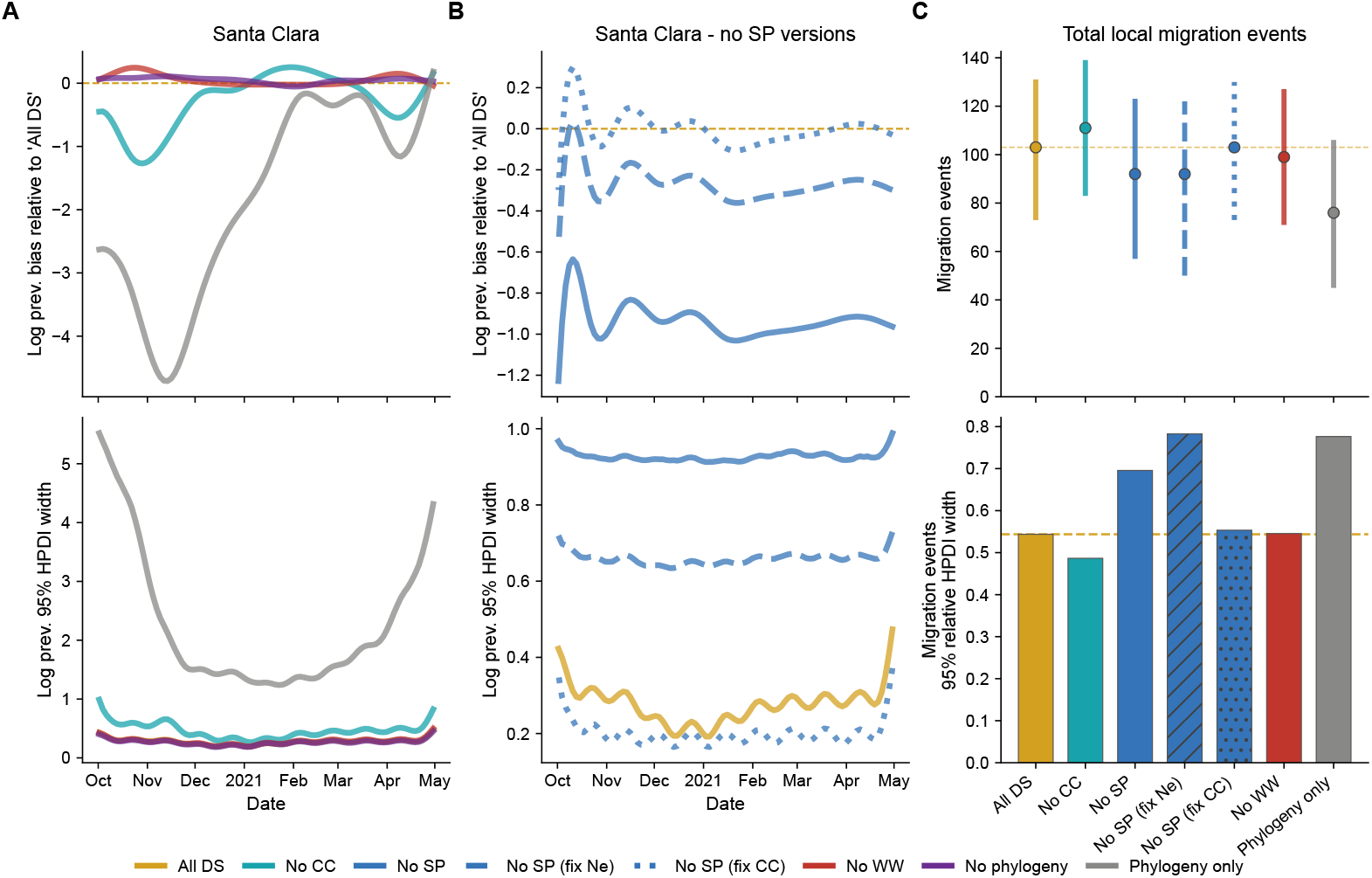
Value of data streams for reconstructing the SARS-CoV-2 winter 2020-21 wave, illustrated for Santa Clara County. Combination of data stream inputs are “All DS”: all data streams included, “No CC”, “No WW”, “No SP”: one of the epidemiological data streams excluded, “No phylogeny”: phylogeny excluded, “Phylogeney”: only phylogeny, i.e. Epsilon variant sequences, provided. A) Bias relative to the “All DS” version and 95% HPD interval (HPDI) width of log prevalence trajectories for Santa Clara county; negative bias values indicate lower inferred prevalence compared to “All DS”. B) As in A) but for the three no-seroprevalence versions: “No SP”, “No SP (fix Ne)”: Ne scaling factor fixed to the median of the “All DS” version, “No SP (fix CC)”: case count scaling factor fixed to the median of the “All DS” version. C) Estimated number of total migration events between counties (whiskers show the 95% HDPI) and their 95% relative HPDI width (width divided by the posterior median); dashed line indicates the median of the “All DS” version for reference. The “No phylogeny” version is omitted since migration events are simply samples from the prior. See also Figures S10, S11 and S12 for additional parameter comparisons.

In contrast, removing case counts led to lower prevalence estimates at the start of the outbreak wave with higher uncertainty (Fig. 4A and Fig. S10A,B), especially at the beginning of the wave in Sacramento and San Francisco, due to less frequent or absent wastewater observations compared to later stages of the outbreak. When case count data was removed, the wastewater standard deviation was estimated to be the lowest of all data combination versions (Fig. S12), likely because the model fitted the prevalence trajectory to the dynamics of wastewater data, requiring less standard deviation, i.e. noise, to explain the wastewater observations. Removing wastewater observations did not change prevalence and migration event estimates (Fig. 4 and Fig. S10). Since both wastewater concentrations and case counts are modeled as direct observations of prevalence, this suggests that when a data stream is very noisy compared to the other data stream, the noisier data does not provide additional information about the prevalence trajectory. In the case of the SARS-CoV-2 Epsilon wave this meant the prevalence trajectory estimate was primarily driven by the case count data. Jointly inferring transmission dynamics on multiple data streams did not improve certainty in the phylogenetic tree. No single data stream seemed to be particularly informative for inferring internal node ages (Fig. S13).

## Discussion

We here introduced MASCOT-DS, a Bayesian phylodynamic approach to accurately reconstruct prevalence trajectories and migration rates in structured populations integrating different data streams: genomic data, case counts, wastewater concentrations and seroprevalence observations. Using simulations and the example of SARS-CoV-2 transmission in the San Francisco Bay Area in Northern California during the Epsilon wave, we show that integrating multiple epidemiological data streams into transmission dynamics inference using MASCOT-DS improves the precision of prevalence trajectories and indirectly, the migration rate estimates.

Quantifying the spatio-temporal transmisison dynamics of SARS-CoV-2 over time between counties in the San Francisco Bay Area during the winter 2020-21 Epsilon wave, we observed frequent transmission between counties and introductions from outside, consistent with the interconnected nature of Bay Area counties and the Bay Area’s role as a domestic and international travel hub [47, 48]. With cases at the beginning being largely driven by local transmission and at the downturn of the wave more so by introductions. At the beginning of the outbreak, the Epsilon variant was more transmissible than co-circulating variants [43], only to be displaced by the Alpha variant in spring 2021 [46]. Part of the observed increase in introduction could also be due modeling the outbreak with fixed instead of time-varying migration rates. Time-varying migration rates could account for changes in transmission due to public health interventions such as increased vaccinations [49]. While we here focused on transmission dynamics between fairly large counties, each with over 0.8 million inhabitants, this method could also be used to quantify transmission between more granular locations, such as neighborhoods, to understand what drives transmission at very local scales [50].

By performing a leave-one-data stream-out analysis, we evaluated the value of each data stream in informing the transmission dynamics estimates. While the phylogeny, informed by genomic pathogen data, is uncertain about the shape and magnitude of the prevalence trajectory in a given location [39, 9, 51], it is the only data stream that can provide information about transmission between locations. We show that we can more precisely estimate migration histories when conditioning on the actual magnitude and dynamics of an outbreak in different locations by integrating epidemiological data streams with the phylogeny. Wastewater concentrations and case counts are both modeled as direct observation processes of the prevalence trajectory, hence, they can stand in for each other, with the less noisy data stream (here case counts) largely informing the posterior estimates. Our simulation study and SARS-CoV-2 analysis suggest that leaving out the noisier data stream has little impact on overall inference. As MASCOT-DS supports any continuous or discrete data type, any additional data stream, such as google search trends for disease symptoms [52], could directly be used as well. Seroprevalence observations are crucial to informing the overall magnitude of an outbreak and genomics can’t replace that using the approach we took here. Without seroprevalence observations, one has to effectively know the under-ascertainment of at least one of the data types and hence, seroprevalence is crucial to quantifying overall disease burden and infectious disease transmission dynamics [53], even one seroprevalence observation paired with frequent case count data during an outbreak can be enough to resolve the scale [32]. We recommend using at least two seroprevalence timepoints in MASCOT-DS for recurring pathogens: one before an anticipated wave to establish baseline immunity from prior infections and one during or after the active or downturn phase of an outbreak to get an estimate of the number of infections accumulated in the course of an outbreak. In addition, we assume that changes in seroprevalence are the result of seroconversion due to infection rather than vaccination. In its current implementation, the model assumes a fully susceptible population at the start of the time period of interest, but it is straightforward to adapt MASCOT-DS to use a prior infection level as input. Alternatively, one can just use seroconversion instead of seroprevalence.

Since we had to use the California wide seroprevalence observations in the absence of county-specific data to inform the magnitude of an outbreak, we effectively assume that an equal fraction of the population was infected in all three locations. To model settings where observations do not neatly overlap with the location boundaries used, MASCOT-DS could be extended to model cumulative numbers across locations instead, which would more accurately reflect uncertainty in infection numbers within locations. This could, for example, be used to model overlapping seroprevalence estimates or instances where sewage catchment areas cover more than one location in the model. However, modeling data streams that only capture a subset of a location can be more challenging. If there are multiple or nested sewersheds in a location of interest [54], the wastewater concentrations could be averaged over wastewater plants for a given observation day. In our SARS-CoV-2 example, Santa Clara reported Anti-N concentrations in four of its sewersheds and we therefore used the mean concentration across sewershed, but ideally would be averaging using a population weighted average across treatment plants.

While investigating the scenarios where we remove different data streams, we did not investigate the scenario where they provide conflicting information. For example, some studies have noted that wastewater and case count trajectories can diverge [55, 16, 56], in part due to different shedding profiles and/or testing strategies that we currently do not model, but that could be added to MASCOT-DS in the future. We also assume that the different data streams are the result of consistent testing proportions over time and currently do not allow to use time varying scaling factors that relate case observations to prevalence.

Our work also shows that using realistic data sizes, case counts are substantially better at capturing the temporal outbreak dynamics than genomics. Case counts in general might be a closer sample of disease incidence, i.e. number of newly infected at a given time, rather than prevalence, i.e. number of infected at a given time, since individuals are more likely to get tested and diagnosed at the first days of symptom onset which would imply that case counts follow a slightly different trajectory compared to the prevalence. However, for infectious diseases that have a relatively short infectious period, such as SARS-CoV-2, the difference between incidence and prevalence trajectory would be small compared to the time scales of the analysis and therefore unlikely to matter. We could adapt MASCOT-DS to accept data streams that inform incidence rather than prevalence.

Future studies could focus on optimizing data collection by running simulations and modifying real-world datasets to test the impact of different measuring frequencies and choices of measurement timepoints for each of the data streams on transmission dynamics estimates. This would allow public health surveillance systems to make cost-effectiveness decisions about investing in the frequency and type of public health surveillance data at different stages of an outbreak [16], in order to achieve a desired level of certainty in parameter estimates of interest. In scenarios where genomic data is sparse, migration rates are poorly informed by phylogeny alone. We anticipate that in these cases, adding epidemiological data streams to inference will not only increase the certainty of the estimates but also speed up convergence, as the posterior landscape may be easier to explore.

In summary, integrating multiple data streams into MASCOT-DS substantially improved certainty in inferring pathogen transmission dynamics in structured populations. We showed that each data stream can contribute distinct and overlapping information about different aspects of transmission dynamics. Compared to traditional structured coalescent methods that estimate effective population sizes and backwards migration rates, MASCOT-DS provides direct estimates of prevalence and forward migration rates, making interpretation much more straightforward. Jointly modeling these data streams also allows users to quantify the value each data stream contributes to informing transmission dynamics metrics of interest. This, in turn, enables cost-effectiveness estimates that can guide decisions about resource allocation at different phases of an outbreak.

## Materials and Methods

### MASCOT-DataStreams implementation

We implemented MASCOT-DataStreams (MASCOT-DS) as a BEAST2 [40] package that extends MASCOT-Skyline [42] to incorporate case counts, wastewater viral concentrations, and seroprevalence surveys in addition to pathogen genomic data. To model nonparametric structured population dynamics through time, we replaced MASCOT-Skyline’s effective population size parameterization *Ne*_*i*_(*t*) with a spline-based representation of prevalence dynamics log *I*_*i*_(*t*) from which *Ne*_*i*_(*t*) is directly derived. This allows case counts, wastewater concentration, and seroprevalence data to each contribute a likelihood term that depends on the same log *I*_*i*_(*t*) trajectory that governs coalescent events in the phylogenetic tree, while keeping MASCOT’s structured coalescent likelihood [41, 42] untouched.

To improve computational efficiency and numerical accuracy, we represent prevalence on a log scale internally. On this scale, true zero prevalence (*I* = 0 is not representable, since *log*(0) = −∞). It can only be approximated by values *log*(*I*) *<* 0, corresponding to 0 *< I <* 1 (fewer than one infected individual). This limitation can be visible in estimates of *log*(*I*) *<* 0 in scenarios where true prevalence is low or zero, which can lead to lower effective sample size (ESS) values as the exact value of *I* between 0 and 1 is not well constrained by the data.

#### Prevalence-spline parameterisation

We parameterise log-prevalence (log *I*_*i*_(*τ*)) in deme *i* as a natural cubic spline *S*(*τ*) anchored at *K* user-specified knot times [*τ*_1_, …, *τ*_*K*_]. Knot times *τ*_*k*_ are given in absolute time backwards from the most recent sample in the phylogenetic tree. Log-prevalence values at knot times, *I*_*i*_(*τ*_*k*_), are directly inferred by MCMC, in between knots log *I*_*i*_(*t*) is interpolated using a natural cubic spline (Fig. S1A). The natural boundary condition means the spline does not extrapolate with curvature beyond the first and last knot, i.e. the log-prevalence trajectory is forced to be constant outside the knot range. Uncertainty of prevalence was higher at knot points compared to the interpolation regions, since during MCMC MASCOT-DS can freely alter the knot values but the variability of prevalence values in between is constrained by the spline interpolation function (Fig. S1B).

The exact number and placement of knot times might take some trial and error, but we recommend to have at least 10 knots, which a knot far enough in the past to include the phylogenetic tree root.

##### Effective transmission rate computation

We derive the effective transmission rate *β*_*i*_(*τ*) at time *τ* analytically using the derivative of the spline 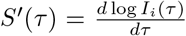 and the becoming uninfectious rate *γ* (Eq. 1). Since the spline prevalence trajectory is expressed backwards in time relative to the most recent samples, we define 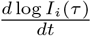 as the change in log-prevalence forwards in time, and 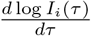 as the change backwards in time, which is equivalent to the derivative of the spline at time *τ*. As an example, Figure S1C shows the transmission rate derived from the estimated prevalence trajectory using simulated data (s. Methods on simulation study).

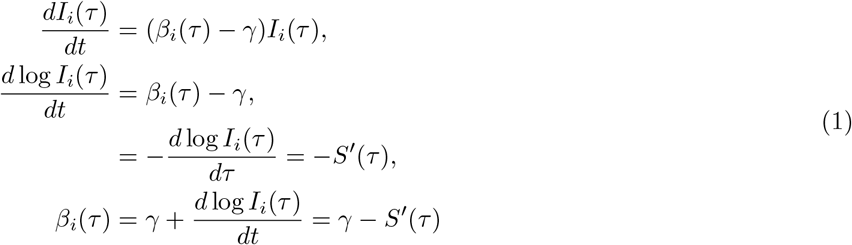

Compared to the transmission rate in an SIR model, *β*_*i*_(*τ*) here can be understood as the effective transmission rate per infectious individual given the proportion of susceptible individuals *S*_*i*_(*τ*)*/N*_*i*_ at time *τ*, 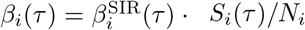. For further details on the transmission rate derivation see Supplementary Methods.

The spline is evaluated on a user-specified dense evaluation grid of timepoints and precomputed prevalence and transmission rates are stored at grid timepoints for efficient lookup during likelihood computations. For a given requested time *τ*, the bordering grid points are found such that *τ*_*left*_ ≤ *τ* ≤ *τ*_*right*_, the log-prevalence and transmission rate at time *τ* are linearly interpolated in between. For query times outside the grid range, the boundary grid-point value is returned. MASCOT-DS uses MCMC to estimate the values of the knot points and then uses the spline interpolation to evaluate the data stream likelihoods at the observation times (Fig. S1A).

##### Hard positivity constraint

We implemented a hard-constraint prior on the prevalence spline of each deme that rejects any MCMC proposal on the prevalence spline that would make any transmission rate on the grid fall below zero due to proposals, i.e. *S*^*′*^(*τ*) *> γ* at any grid point.

#### MASCOT integration

We use the prevalence trajectories and forward migration rates to derive the parameters required by the structured coalescent likelihood of MASCOT, namely the time-varying effective population sizes *Ne*(*t*) for each deme and the backward migration rates between demes. Due to MASCOT-DS being a package in the BEAST2 ecosystem, the phylogenetic tree can either directly be provided or estimated from user-provided pathogen sequences.

##### Effective population size computation

We calculate the effective population size *Ne*_*i*_(*τ*) at time *τ* analytically using the prevalence *I*_*i*_(*τ*) and the transmission rate *β*_*i*_(*τ*) as derived by [39] (Eq. 2).

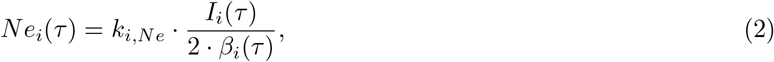

We introduce a deme-specific scaling factor *k*_*i,Ne*_ that can be estimated through MCMC to allow for non-homogeneous transmission dynamics, such as superspreading (*k*_*i,Ne*_ *<* 1) [9, 51]. As an example, Figure S1D shows the *Ne* derived from the estimated prevalence and transmission rate trajectory using simulated data (s. Methods on simulation study).

##### Backward migration rate computation

In MASCOT-DS we estimate the forward migration rate 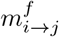 which estimates the rate of a lineage moving from deme *i* to deme *j* forwards in time. However, the structured coalescent likelihood requires the backward migration rate 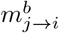 which estimates the rate of a lineage moving from deme *j* to deme *i* backwards in time. Forward and backward migration rates are related by Eq. 3.

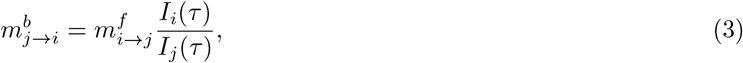

All these components constitute the structured coalescent likelihood *MASCOT* (*D*_*T*_|*Ne, m*), where *D*_*T*_ refers to the pathogen phylogeny, which describes the phylogenetic trees estimated from pathogen genomic data.

#### Data stream likelihoods

We introduced three novel likelihoods to incorporate epidemiological data, such as case counts *D*_*cc*_, wastewater viral concentrations *D*_*ww*_, and seroprevalence surveys *D*_*sp*_, into MASCOT-DS’ transmission dynamics inference. MASCOT-DS supports two types of data that are direct observations of prevalence: a likelihood for discrete, non-negative data such as case counts, and a likelihood for continuous, strictly-positive data such as wastewater viral concentrations. The third likelihood, described below, models seroprevalence data, which reflects cumulative incidence built up over time rather than prevalence at a single moment, and therefore requires a different way of linking the data to the underlying transmission dynamics. Table 1 and Figure 1A summarize the likelihoods and parameters for each of the four data streams.

##### Case count likelihood *P* (*D*_*cc*_|*µ, α*_*cc*_)

We assume case counts *cc*_*i*_(*t*) in deme *i* at time *τ* to be distributed according to a negative binomial distribution, aka Gamma-Poisson mixture, with a dispersion parameter *α*_*cc*_ and the mean *µ* being informed prevalence *I*_*i*_(*τ*) multiplied by a deme-specific scaling factor *k*_*cc,i*_ (Eq. 4).

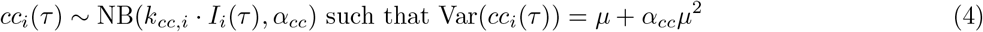

The scaling factor *k*_*cc,i*_ accounts for reporting fraction of reported cases to the latent unobserved prevalence *I*_*i*_(*τ*). The dispersion parameter *α*_*cc*_ describes the noisiness of the case count data. Both parameters can be fixed by the user or estimated through MCMC sampling. This likelihood can accept any discrete, non-negative data stream.

##### Wastewater viral concentration likelihood 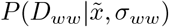

We assume the wastewater viral concentrations *ww*_*i*_(*τ*) in deme *i* at time *τ* to be distributed according to a log-normal distribution with a standard deviation *σ*_*ww*_ and the median 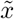 being the proportion of infected individuals in the population *I*_*i*_(*τ*)*/N*_*i*_ multiplied by a deme-specific scaling factor *k*_*ww,i*_ (Eq. 5). We calculate the mean of the log-normal distribution (in log space) as 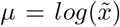. Log-normal distributions are frequently used to model properties of wastewater surveillance data [27, 57, 58].

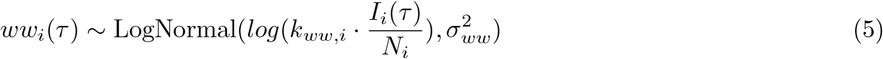

The scaling factor *k*_*ww,i*_ accounts for deme-specific factors influencing the relationship between proportion of infected individuals and wastewater viral concentrations. The standard deviation *σ*_*ww*_ describes the noisiness of the wastewater viral concentration data. Both parameters can be fixed by the user or estimated through MCMC sampling. This likelihood can accept any continuous, strictly positive data stream.

We assume wastewater viral concentrations to be reported as normalised to pepper mild mottle virus (PMMoV) concentrations to control for variations in fecal content and dilution across samples.

##### Seroprevalence likelihood *P* (*D*_*sp*_ | *n*_*tested*_, *p*)

We assume the number of seropositive individuals *n*_*ab*+,*i*_(*τ*) out of *n*_*tested,i*_(*τ*) tested in deme *i* at time *τ* to be distributed according to a binomial distribution with probability *p*_*i*_(*τ*), reflecting the proportion of cumulative infected people in the tested population (Eq. 6).

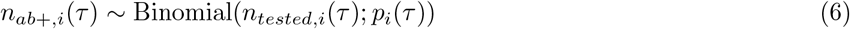

We derive the seropositivity probability *p*_*i*_(*τ*) from the cumulative incidence *C*_*i*_(*τ*_*K*_, *τ*) as 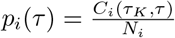. MASCOT-DS expects 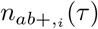 and *n*_*tested,i*_(*τ*) as inputs and rounds them to the nearest integer. If a user assumed that the seroprevalence observations were obtained from a subpopulation with a different hazard of infection compared to the general population, a scaling factor can be introduced (Supplementary Methods). However, we note that tight, well-informed priors should be added with this scaling factor to maintain the ability to accurately inform the overall magnitude of prevalence and we can hardly imagine a scenario where this would be reasonably possible.

MASCOT-DS evaluates the joint likelihood of the structured coalescent likelihood, the data stream likelihoods and any priors (Eq. 7) using the BEAST2 MCMC inference framework.

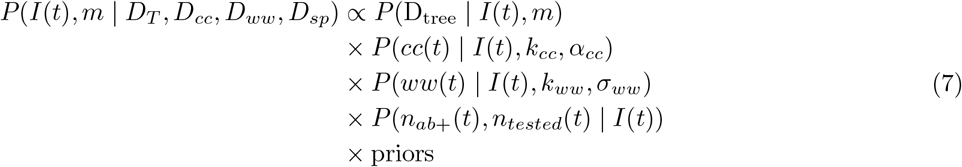

MASCOT-DS users can export prevalence, cumulative incidence, transmission rates and effective population sizes for user-specified timepoints along the evaluation grid per MCMC step via the common BEAST2 logging infrastructure.

### SIR simulation study

The simulation study was designed to evaluate the performance of MASCOT-DS in recovering structured transmission dynamics from simulated data streams. We simulated outbreaks in two demes under the SIR model using ReMASTER (v2.7.2) [59]. In total we performed 100 independent simulations, for details on the settings we used in ReMASTER see Supplementary Methods. The goal of our simulation study was to approximate a well-calibrated simulation study [60]. The study is an approximation, since in our case the SIR model used in the simulation differed from the coalescent model used during inference.

#### Data stream simulation

We simulated case counts, wastewater viral concentrations, and seroprevalence from the infected population trajectories *I*_*i*_(*t*) obtained from the ReMASTER simulation using custom python scripts. We restricted data stream simulations to timepoints before the most recent sampled sequence in the phylogenetic tree of a given simulation.

##### Case count simulation

For each simulation and deme, we sampled case count observations *cc*_*i*_(*t*) at daily intervals. At a given time *t* the case counts were sampled from a negative binomial (Gamma-Poisson mixture) distribution with mean *µ*_*i*_(*t*) = *I*_*i*_(*t*) · *k*_*i,cc*_ and dispersion parameter *α*_*cc*_, such that the variance of case count observations followed *µ*_*i*_(*t*) + *α*_*cc*_ · *µ*_*i*_(*t*)^2^ (Eq. 8). For each deme, the scaling factor *k*_*i,cc*_ was independently sampled from a log-normal distribution *k*_*i,cc*_ ~ LogNormal(−3, 0.5). The dispersion parameter was the same for both demes and was sampled from a log-normal distribution *α*_*cc*_ ~ LogNormal(−1, 0.5). This was done separately for each simulation.

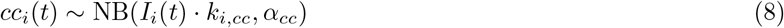

##### Wastewater viral concentration simulation

For each simulation and deme, we sampled a series of wastewater viral concentration observations *ww*_*i*_(*t*) at daily intervals. At a given time *t*, we sampled *ww*_*i*_(*t*) from a lognormal distribution with standard deviation *σ*_*ww*_ and the median 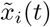 being the proportion of infected individuals in the population *I*_*i*_(*t*)*/N*_*i*_ scaled by a scaling factor *k*_*i,ww*_ (Eq. 9). For each deme, the scaling factor *k*_*i,ww*_ was sampled from a log-normal distribution *k*_*i,ww*_ ~ LogNormal(4.5, 0.5). The standard deviation *σ*_*ww*_ was sampled from a log-normal distribution *σ*_*ww*_ ~ LogNormal(−0.7, 0.3) and was shared across both demes. Since at *I*_*i*_(*t*) = 0 the wastewater viral concentration is undefined under the above simulation regime, we didn’t report wastewater observations for timepoints where *I*_*i*_(*t*) = 0.

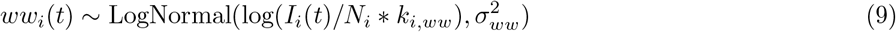

##### Seroprevalence simulation

For each simulation and deme, we generated three seroprevalence observations at distinct phases of the outbreak: beginning, middle, and end. At time *t*, we first defined the probability of having been infected as the proportion of cumulatively infected individuals *C*_*i*_(*t*) in the population as

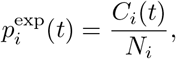

where we clipped 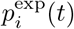 to [10^−16^, 1 − 10^−16^] for numerical stability. We then simulated seroprevalence *sp*_*i*_(*t*) as the fraction of individuals testing positive for antibodies *n*_*ab*+,*i*_(*t*) out of a number of individuals tested *n*_*tested,i*_(*t*), which was sampled from a Uniform distribution (Eq. 10).

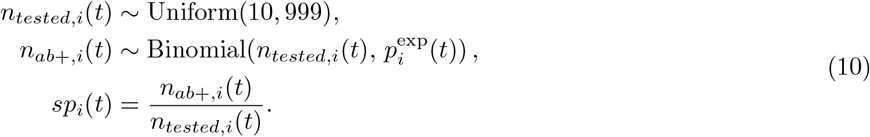

We sampled one serosurvey observation during the beginning of the outbreak (within the first 20% of the outbreak time range, where 100% is the timepoint of the most recent sample), during the middle (40% − 60%), and one during the downturn of the outbreak (80% − 100%) with the exact timepoint being randomly chosen.

For details on how we ran MCMC inference under the MASCOT-DS model on the simulation data within BEAST2 see Supplementary Methods.

#### Posterior predictive check

To assess whether the fitted model reproduced the observed wastewater concentration data, we performed a posterior predictive check (PPC) for each simulated dataset. For each of the 100 simulations and each wastewater observation timepoint, we drew 1000 wastewater concentration values using the posterior prevalence trajectory, wastewater scaling factor and standard deviation posterior distributions according to the wastewater log-normal likelihood function. This produced, for every real observation, a full posterior predictive distribution of replicate concentrations from which we could calculate the fraction of true observations contained in the 95% posterior predictive interval for each simulation.

### SARS-CoV-2 Epsilon wave in the Bay Area

We chose October 1st, 2020 until May 1st, 2021 as the time period of interest, which followed the SARS-CoV-2 winter 2020-21 wave in the San Francisco Bay Area. Within that period, the share of the Epsilon variant among all SARS-CoV-2 sequences in California in GISAID [61] rose from 2.1% in the week of September 28, 2020 to its peak of 60.5% during the week of February 15, 2021 with a sharp decline to 8.0% at the end of the our studied time period [46]. Epidemiological data (case counts, wastewater concentrations and seroprevalence observations) collected within the time period of interest was used in this study.

#### Sequence data acquisition and processing

We downloaded all SARS-CoV-2 Epsilon sequences in North America from the GISAID database using the variant identifiers B.1.427 and B.1.429 and required the host to be human. We excluded low coverage sequences and only included sequences with complete genome coverage and complete date information. In addition, we only considered sequences collected before May 1st 2021 to focus on the winter 2020-21 wave in the San Francisco Bay Area. Based on overall data availability, we designated three counties as our demes of interest: San Francisco, Sacramento and Santa Clara, as all of them had a sufficient number of sequences available as well as consistent case count and wastewater reporting during the winter 2020-21 wave.

We aimed to construct a 1000 sequence multiple sequence alignment by randomly selecting 310 Epsilon sequences in each county and 70 background sequences randomly chosen across the US to serve as a background pool from which lineages could migrate into the counties of interest to estimate the tree structure for the three local counties and to estimate introductions into the counties from outside. The number of 70 background sequences was arbitrarily chosen. We aligned sequences using MAFFT v7.490 [62] in auto mode with 4 threads. Ambiguous bases (N) were replaced with gaps prior to trimming gaps with trimAl v1.5 [63] using a gap threshold of 0.9, which removed columns in the alignment covered by less than 90% of the sequences. Using IQ-TREE v3.0.1 [64], we inferred a maximum-likelihood tree that we then time-calibrated with TreeTime v0.11.4 [65] to detect molecular clock outliers. One San-Francisco sequence was identified as an outlier and removed from the alignment. The final multiple sequence alignment contained 999 sequences and had a length of 29186 bases.

All genome sequences used in this study and associated metadata can be accessed through the EPI SET 260725qm identifier (https://doi.org/10.55876/gis8.260725qm, Supplementary Material).

#### Data stream acquisition and preparation

##### Case count data

We obtained daily case counts for each county of interest from the California Department of Public Health (https://data.chhs.ca.gov/dataset/covid-19-time-series-metrics-by-county-and-state/resource/046cdd2b-31e5-4d34-9ed3-b48cdbc4be7a, downloaded: 03/02/2026). Rows with masked counts and weekend dates (Saturday-Sunday) were excluded, since weekend reporting was systematically lower than weekday reporting.

##### Wastewater viral concentration data

We obtained daily wastewater SARS-CoV-2 concentrations from the California Sewage Coronavirus Alert Network (SCAN) network (https://data.chhs.ca.gov/dataset/wastewater-surveillance-data-california downloaded: 03/02/2026,[66]). These data were collected as part of the WastewaterSCAN / SCAN project, a partnership between Stanford University, Emory University, and Verily funded philanthropically through a gift to Stanford University. We filtered for records targeting the SARS-CoV-2 N gene and used Pepper mild mottle virus (PMMoV) as the fecal indicator to normalise the SARS-CoV-2 N concentrations to the PMMoV concentrations and multiplying by 1, 000, 000 to match the methodology described on the WastewaterSCAN dashboard [5]. We removed wastewater concentration measurements of 0 since the wastewater likelihood (log-normal) in MASCOT-DS can’t handle concentrations of 0. Santa Clara county reported concentrations across four wastewater treatment plants, so we averaged concentrations across all plants for a given collection date.

##### Seroprevalence survey data

We obtained statewide anti-nucleocapsid (anti-N) seroprevalence values from the NCHS Nationwide Commercial Laboratory Seroprevalence Survey (https://data.cdc.gov/Laboratory-Surveillance/Nationwide-Commercial-Laboratory-Seroprevalence-Su/d2tw-32xv/data_preview, downloaded: 03/02/2026). We used the survey results in California from survey rounds 1-30 using the all age group measurements. The data directly provided the number of individuals tested *n*_*tested*_ and the rate of seropositivity for Anti-N antibodies for all ages. We used the Anti-N data, which measures prior infection rather than response to vaccination. To obtain the number of individuals tested positive for antibodies *n*_*ab*+_, we multiplied the number of tested individuals by the rate of seropositivity. The measurements were provided for a date range, we picked the last date of the range as the timepoint of measurement.

Since seroprevalence surveys were conducted at the state level, we used the California wide seroprevalence data as stand-in seroprevalence observations for each individual county, assuming a similar level and trajectory of infection between counties and across California.

For details on how we ran MCMC inference under the MASCOT-DS model on the SARS-CoV-2 data within BEAST2 see Supplementary Methods.

#### Calculating percent introductions in new cases

We calculated the total number of new cases in a given deme as the total transmission rate multiplied by the prevalence in a given deme *β*_*i*_(*t*) · *I*_*i*_(*t*). We computed the number of new cases due to introductions from other demes including the outside deme as the sum of the product of migration rates and prevalences across other demes 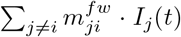. This yields the percentage of new cases due to introductions as a percentage of the total new cases in a given deme 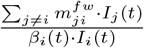. Note, that this is a very strict definition of cases due to introductions as it does not attribute secondary cases seeded by an introduction into the percentage.

### Value of information of different data streams

For simulation study and real world application to the SARS-CoV-2 Epsilon wave in the Bay Area, we gave MASCOT-DS different combinations of data streams as input. We tested following combinations: removing case counts (“no CC”), wastewater (“no WW”), seroprevalence (“no SP”) and the phylogeny, i.e. prevalence and migration rates estimates were not evaluated on the tree (“no phylogeny”), this meant migration rates were simply sampled from the prior. In addition, we also tested the results when just providing SARS-CoV-2 Epsilon sequences as input, i.e. only the phylogenetic tree was used to infer the prevalence and migration rates, but all epidemiological data streams were removed (“Phylogeny only”).

In the case of the simulation study, we subtracted the ground truth value from the median of the posterior samples of each parameter for each data stream combination version to assess bias, we calculated the relative bias by further dividing by the ground truth value. In the case of the SARS-CoV-2 San Francisco Bay Area application, we subtracted the posterior median value of the all data streams model (“All DS”) from the median of posterior samples of each parameter for each data stream combination version to assess bias, we calculated the relative bias by further dividing by the “All DS” posterior median value.

In the simulation study, both demes had complete data streams available and the Ne scaling factor was fixed to 1 which mean that migration rates were directly comparable between data stream combination versions. In contrast, in the SARS-CoV-2 Bay Area application there was some ambiguity between Ne and migration rate estimates, as the model could explain the phylogeny by either changing the Ne or the migration rate. This was particularly relevant for the outside deme with no calibrating epidemiological data stream, so its prevalence magnitude was only weakly constrained. Because the effective population size of a sparsely sampled deme trades off against its migration rates in the structured coalescent likelihood, the migration rates are not reliably identified and thus not comparable across data stream combination versions. The number of migration events, in contrast, is estimated directly from the reconstructed genealogy and is the more meaningful quantity to compare.

To assess changes in uncertainty, we compared the width of the 95% HPD intervals between the different data stream combination versions. We calculated the relative width by dividing further by the median of the corresponding posterior samples. In the case of the SARS-CoV-2 San Francisco Bay Area application when removing seroprevalence observations, we also tested the value of fixing the effective population size scaling factor or the case count scaling factors to the median values of the corresponding posterior in the “All DS” model to address potential non-identifiability of the scaling factors *k*_*Ne*_, *k*_*cc*_, *k*_*ww*_ when seroprevalence was removed.

For all data stream combination versions, we ran the inference as described in the ‘SIR simulation study’ and ‘SARS-CoV-2 Epsilon wave in the Bay Area’ methods section, respectively. In the case of the “Phylogeny only” version, we also applied a Normal(0, 1) smoothing prior on successive prevalence knot values of each county’s prevalence trajectory to help regularise the prevalence trajectory. In the case of the “no phylogeny” version, we had the structured coalescent likelihood return a log-likelihood of 0, effectively removing any evaluation on migration rates and prevalence estimates based on the phylogeny, we also removed all tree operators in this version. In addition, we fixed the outside deme prevalence to a log-prevalence of 6 and did not estimate it, since without estimating the tree, there was nothing informing the outside deme’s prevalence.

#### Computational cost of adding multiple data streams

We quantified the computational cost of using multiple data streams for parameter estimation by comparing, across data stream combinations, (i) the number of MCMC samples and wall-clock time required to reach stationarity, and (ii) the wall-clock time subsequently required to reach adequate mixing (effective sample size, ESS ≥ 200). We performed this calculation on results from simulated data, as this gave us sufficient sample sizes for comparison. Each metric was computed independently for 10 simulations x six data stream input combinations x 3 replicate-seed combination (180 runs). For details on how we calculated these metrics see Supplementary Methods.

## Data Availability

All scripts and instructions on reproducing the results of this study can be found at https://github.com/Pweidemueller/MASCOT-DS_materials.

https://github.com/Pweidemueller/Mascot_datastreams

https://github.com/Pweidemueller/MASCOT-DS_materials

## Data availability

All scripts to generate BEAST2 XMLs from input data, analyze and plot results, are available on GitHub at https://github.com/Pweidemueller/MASCOT-DS_materials/. The source code for the BEAST2 package MASCOT-DS itself is available on GitHub at https://github.com/Pweidemueller/Mascot_datastreams.

## Acknowledgments

This study was funded by a UC Noyce initiative award. PHW, IRB, and NFM are funded in part by the UC Noyce initiative award. We thank Joyce Lee for providing valuable feedback on the manuscript. We thank Elana Chan for an insightful discussion on different properties of wastewater data. We gratefully acknowledge all data contributors, i.e., the Authors and their Originating laboratories responsible for obtaining the specimens, and their Submitting laboratories for generating the genetic sequence and metadata and sharing via the GISAID Initiative, on which this research is based.

## Supplementary Figures

**Supplementary Figure S1:**
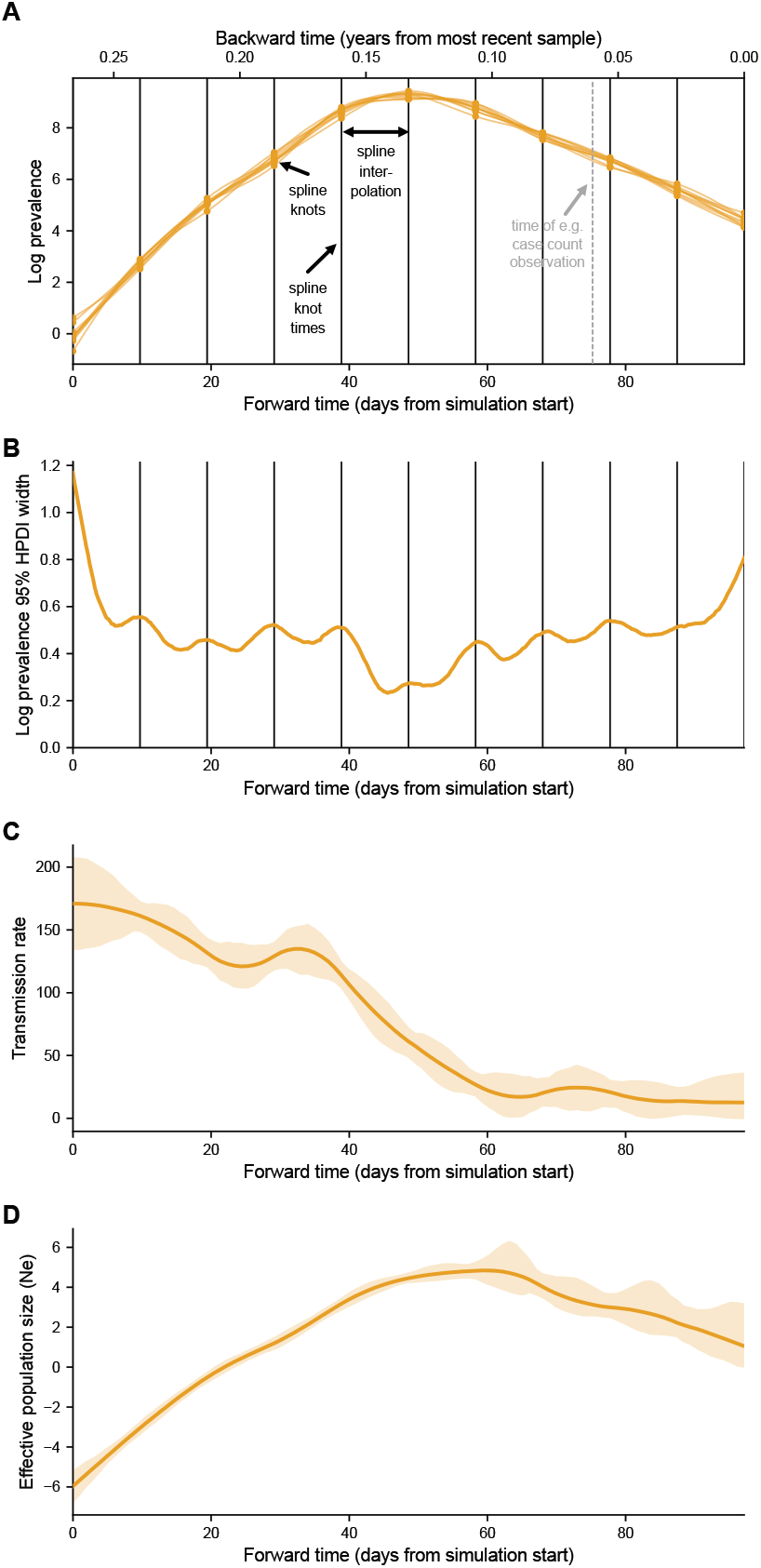
Example of prevalence trajectory spline interpolation and effective population size calculation. A) Ten samples from the posterior of the inferred log prevalence trajectory (orange lines) of one example SIR outbreak simulation. MASCOT-DS estimates the log prevalence values at knot points (orange dots) at given times (black vertical lines) and interpolates the prevalence in between using a cubic spline. Knot times were provided to MASCOT-DS backwards in time (*τ*) relative to the most recent genomic sample. The dashed, gray line shows the time of an example case count observation, MASCOT-DS takes the prevalence at this timepoint to evaluate the data stream likelihood for this observation. B) 95% HPD interval of the log prevalence trajectory of this example simulation. Black vertical lines denote the spline knot times. C) Transmission rate calculated from the log prevalence trajectory posterior of this example simulation. The orange shaded area spans the 95% HPD interval, the orange solid line shows the median. D) Effective population size *Ne* calculated from the log prevalence and transmission rate trajectories of this example simulation. The orange shaded area spans the 95% HPD interval, the orange solid line shows the median.

**Supplementary Figure S2:**
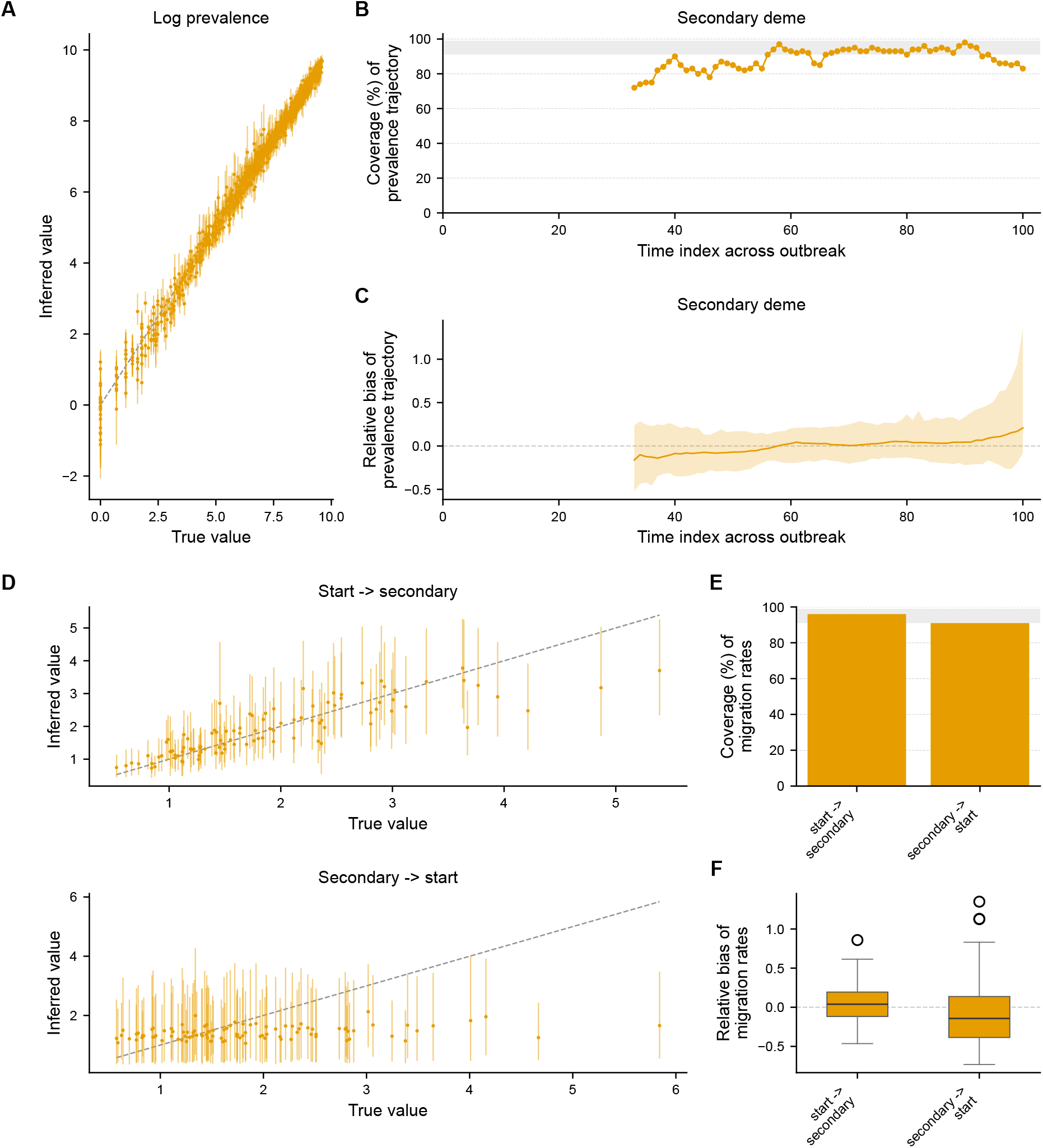
MASCOT-DS validation using 100 two-deme SIR simulations. A) Scatter plot of the inferred log prevalence values against the true log-transformed prevalence values for the secondary deme across 100 simulations. Each dot is the median of the posterior of one of 11 knot points of the prevalence trajectory of a given simulation, whiskers span the 95% HPD interval. Since the time index at which the outbreak arrived in the secondary deme from the start deme differed between simulations, coverage is only shown for time points at which there was at least one infected across all 100 simulations. B) Coverage of true prevalence values in the 95% HPD interval of the estimated prevalence trajectory across all simulations. Grey band indicates desired 91-99% coverage. C) Relative bias of prevalence estimates relative to the true prevalence values. For each simulation, the true value was subtracted from the median of the posterior distribution and divided by the true value. The line shows the median bias across simulations, shaded area the 2.5 and 97.5 percentiles. D) Scatter plots of the inferred migration rates against the true migration rates. E) Coverage of the true migration rates across 100 simulations. F) Relative bias of migration rate estimation relative to the true migration rates.

**Supplementary Figure S3:**
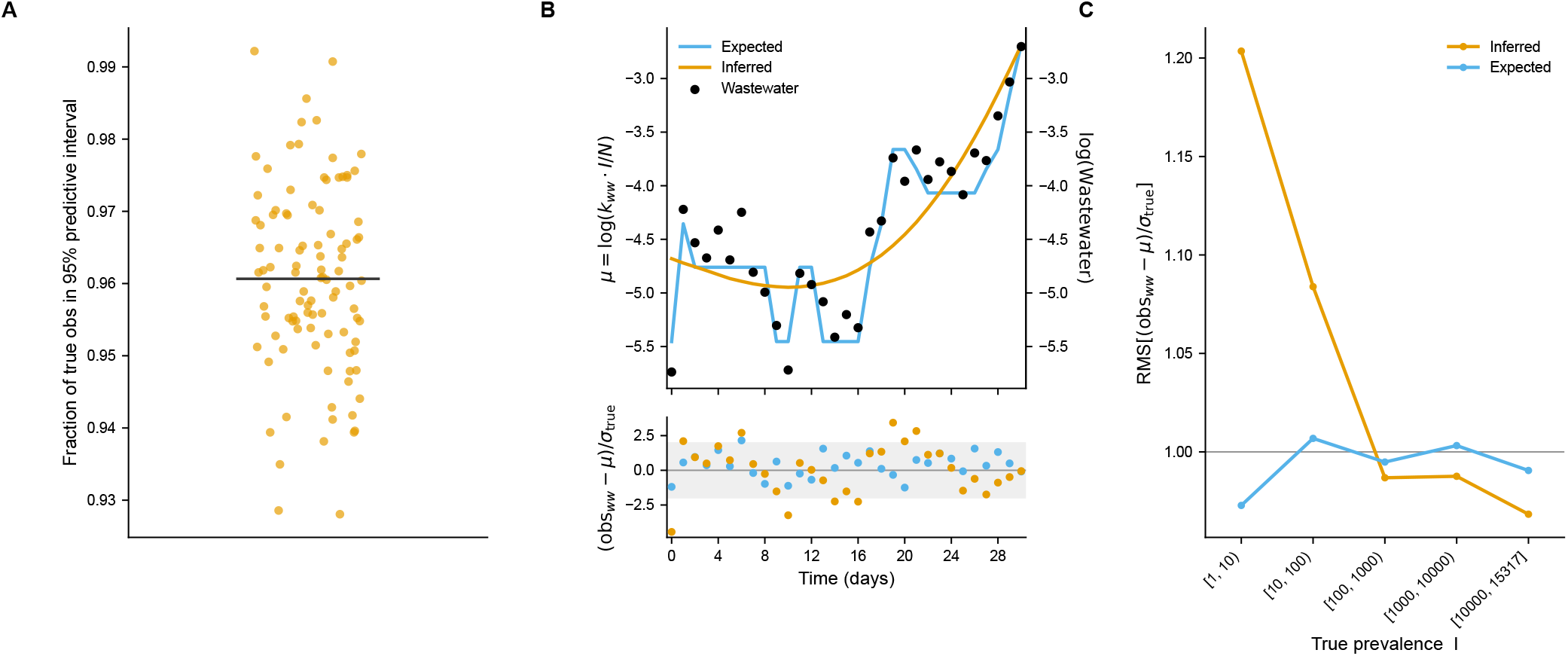
Posterior predictive check of MASCOT-DS’ fit of simulated wastewater data. A) Posterior-predictive coverage across the 100 simulations. Each dot is one simulation; the *y*-axis is the fraction of its true wastewater observations (obs) falling within the 95% posterior-predictive interval (median across simulation is shown as the black line). B) Example of prevalence misfit during the early, low-prevalence phase of an outbreak for one simulation. Top panel shows the expected (blue) and inferred (orange) *µ* of the wastewater log-normal distribution at each wastewater observation (black dots) timepoint. The bottom panel shows the corresponding standardized residuals (obs_*ww*_ − *µ*)*/σ*_true_ against the inferred and expected *µ*. The grey band covers the 95% highest density interval of a standard normal distribution. Residuals are expected to fall in this range if the model would be able to perfectly fit the prevalence trajectory. C) Root-mean-square (RMS) of the standardized residuals of the inferred and expected *µ* standardized to true observation noise *σ*_true_, pooled across all simulations and binned by true prevalence counts *I*. Grey line shows RMS at 1, which indicates no bias in residuals relative to *σ*_true_.

**Supplementary Figure S4:**
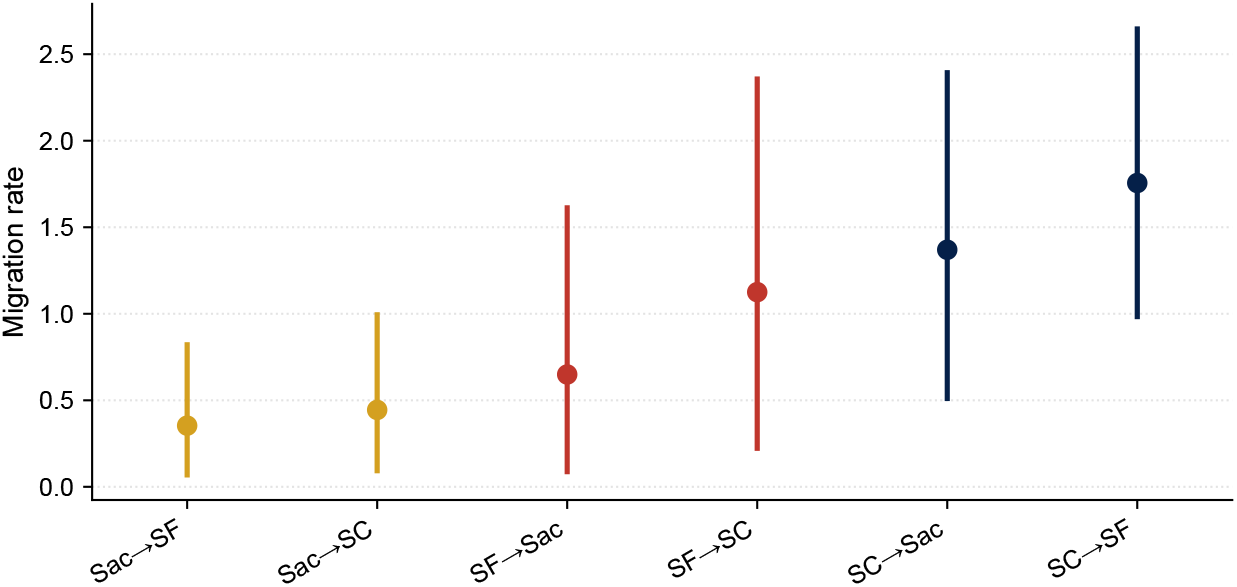
Migration rates between Bay Area counties Sac=Sacramento, SC=Santa Clara, SF=San Francisco) during the SARS-CoV-2 Epsilon wave. Dots are the median of the posterior distribution, whiskers span the 95% HDP interval.

**Supplementary Figure S5:**
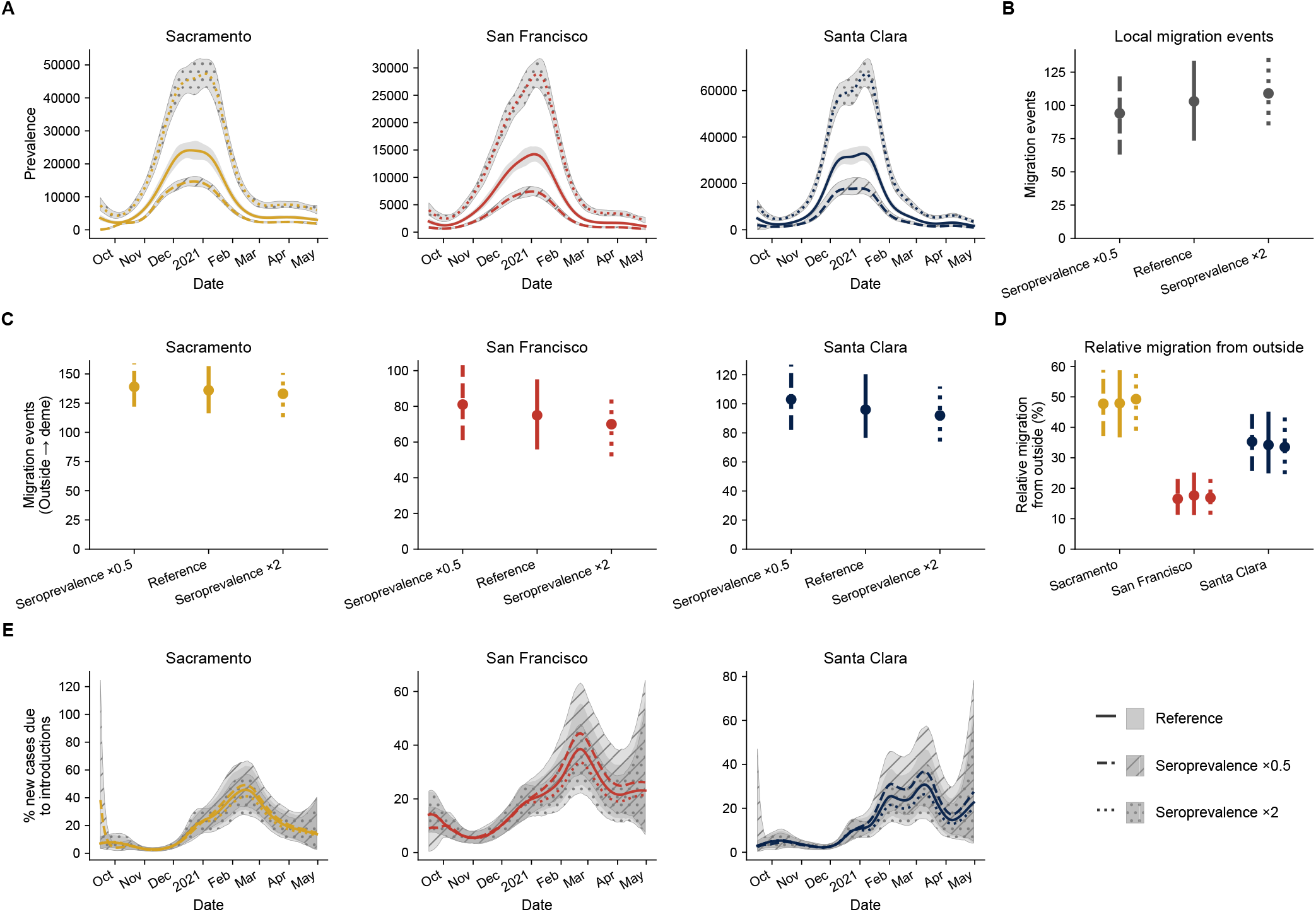
Effect of different seroprevalence value scales on informing various transmission dynamics metrics in the SARS-CoV-2 2020-21 wave. We artificially scaled seroprevalence values by 2 (dotted) or 0.5 (dashed) and provided all other data streams unaltered to MASCOT-DS. A) Prevalence estimates for the three San Francisco Bay Area counties. The truly reported seroprevalence observations serve as a reference. Colored, solid lines are the median of the posterior distribution, grey shaded areas with different hatches denote the 95% HPD interval. B) Relative migration strength from outside the Bay Area into each country. Dots are the median of the posterior distribution, whiskers denote the 95% HPD interval. C) Number of estimated migration events from the outside deme into each county. D) Total estimated local migration events between counties. E) Estimated percent of new cases due to introductions into a given county.

**Supplementary Figure S6:**
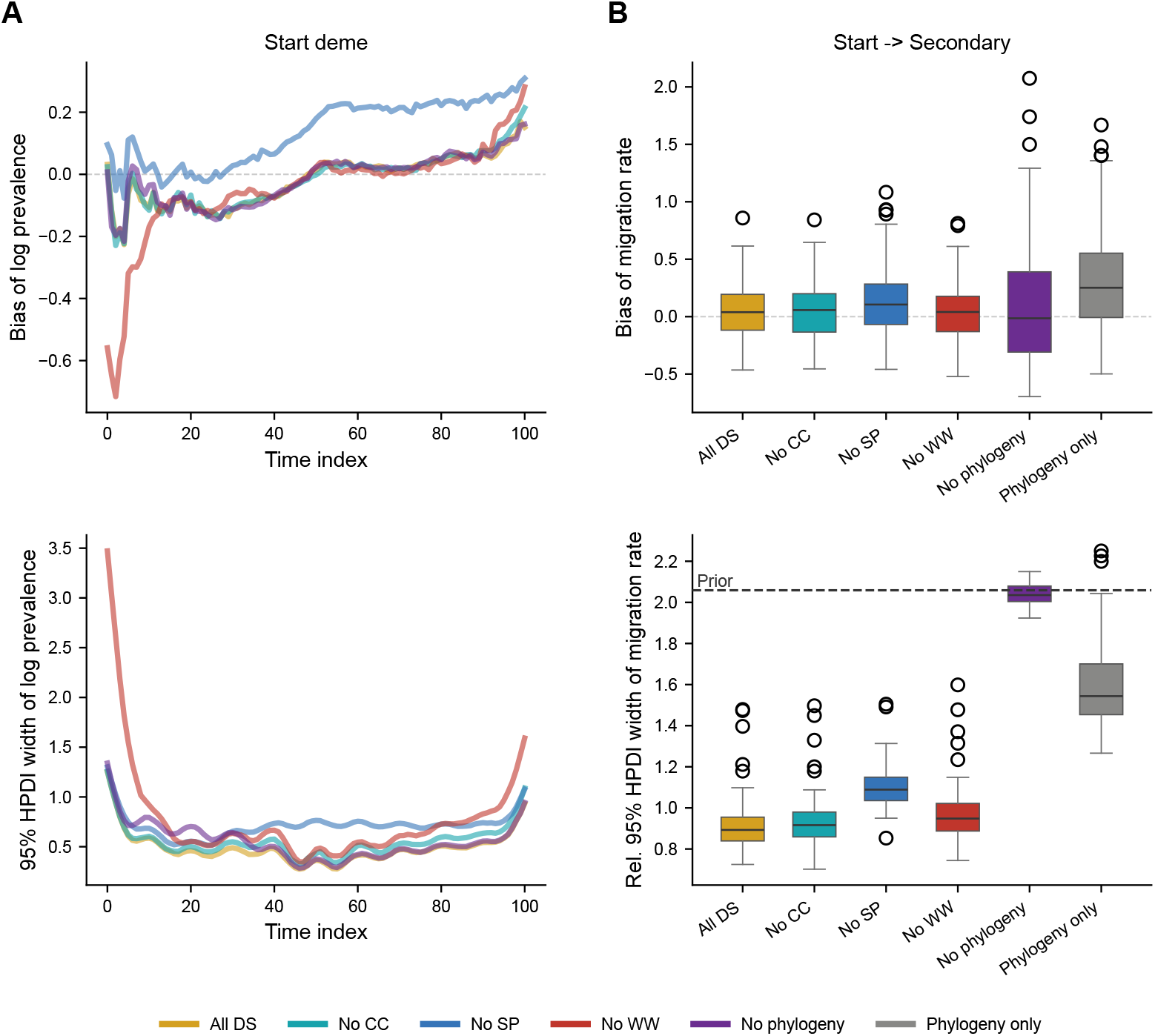
Value of information of each data stream in the simulation study. MASCOT-DS inference estimates when individual data streams were excluded, compared to the true (simulated) values. Estimates for the starting deme are shown here; estimates for the secondary deme are shown in Fig. S7. “All DS”: all data streams included, “No CC”, “No WW”, “No SP”: one of the epidemiological data streams excluded, “No phylogeny”: phylogeny excluded, “Phylogeny only”: only phylogeny, i.e. phylogenetic tree, provided. A) Bias and 95% HPD interval (HPDI) width of log prevalence trajectories for different data stream input combinations. Lines show the median bias and HPDI width across simulations. Time index 0 = outbreak start, increasing forward in time; “Phylogeny only” is omitted here as its uncertainty dominated the axis (shown in Fig. S7). Dashed line marks zero bias. B) Relative bias and 95% HPDI width of the start-to-secondary dme migration rate. Boxplots show the spread across simulations (box = IQR, line = median, whiskers = 1.5 *×* IQR, points = outliers). Grey, dashed line marks zero bias; black, dashed line marks the 95% highest density interval of the prior distribution of the migration rates.

**Supplementary Figure S7:**
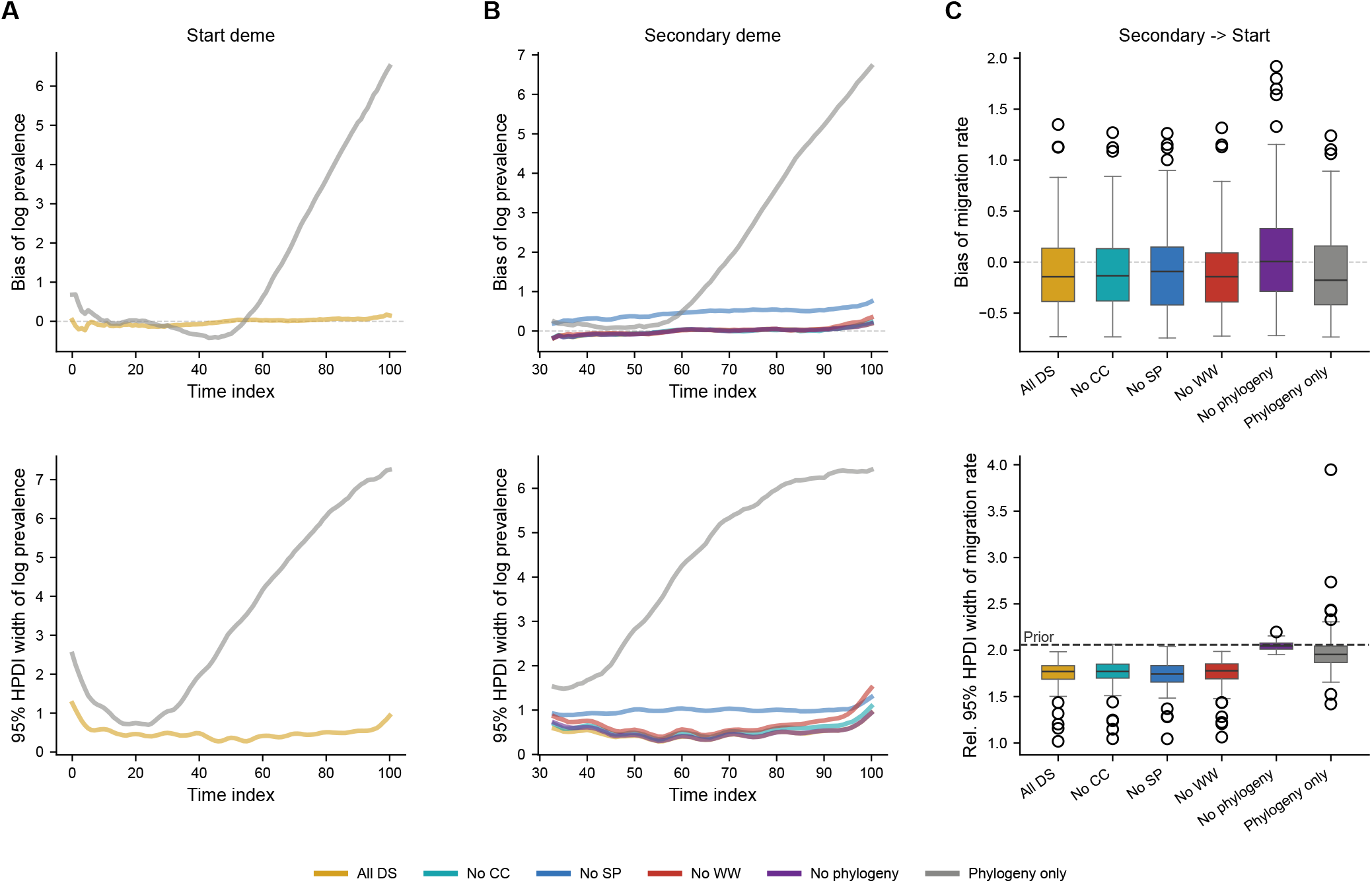
Value of information of each data stream in the simulation study. Plots show inference estimates of MASCOT-DS when certain data streams were excluded. Estimates are compared to the true values based on the simulated data. A) Bias and 95% HPD interval (HPDI) width of log prevalence trajectories of the start deme when only the phylogeny was provided as input compared to the “All DS” model. Lines show the median bias and HPDI width across simulations. Time index 0 = outbreak start, increasing forward in time B) Bias and 95% HPD interval (HPDI) width of log prevalence trajectories of the secondary deme for different data stream input combinations. Only time indices where prevalence was *>* 0 in all simulations are shown. C) Relative bias and 95% HPD width of the secondary → start migration rate. Boxplots show the spread across simulations (box = IQR, line = median, whiskers = 1.5*×*IQR, points = outliers). The grey, dashed line marks zero bias; the black, dashed line marks the 95% highest density interval of the prior distribution of the migration rates.

**Supplementary Figure S8:**
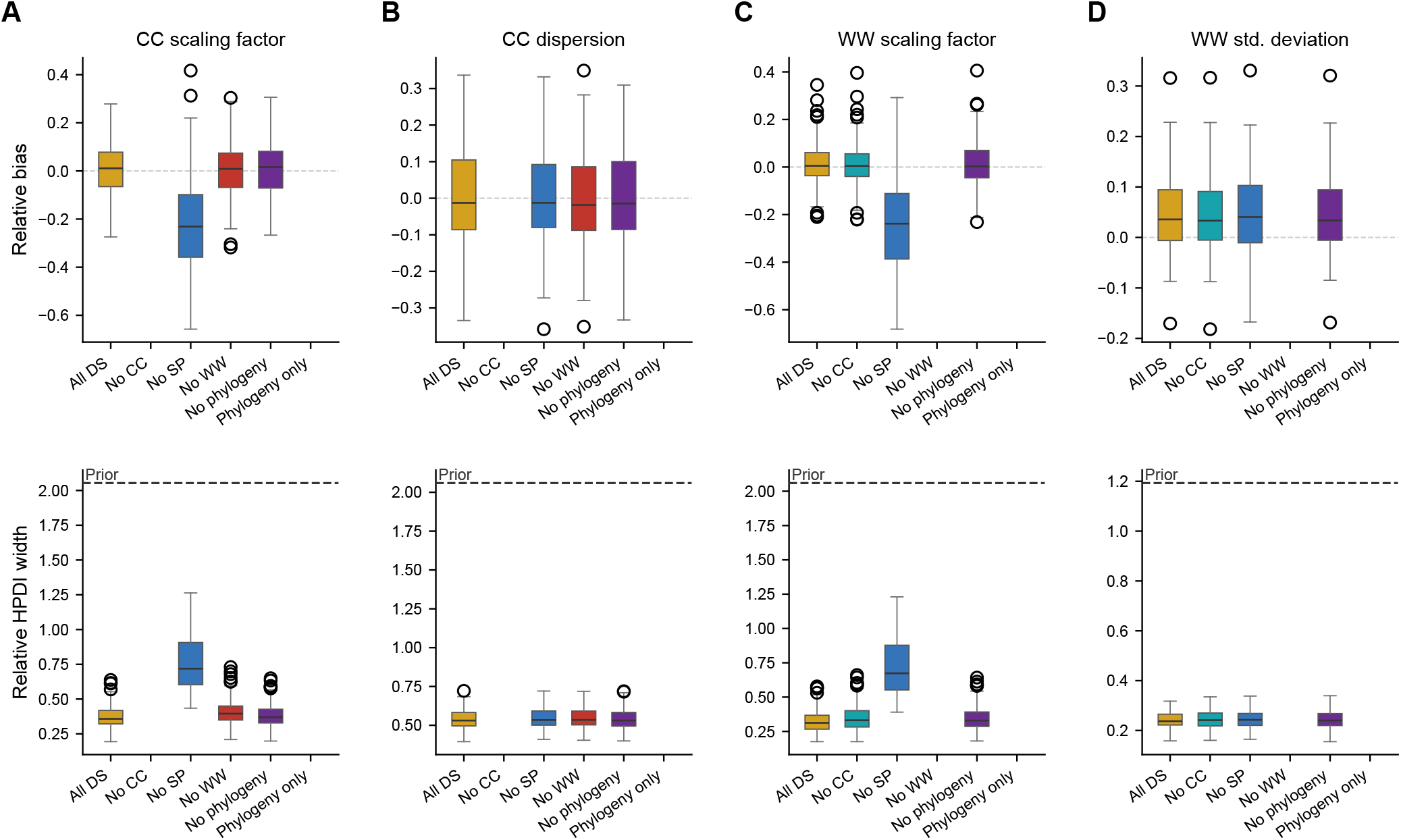
Recovery of data stream likelihood parameters when certain data streams were excluded as input into MASCOT-DS in the simulation study. Estimates are compared to the true values based on the simulated data. When only the phylogeny was given as input, no case count and wastewater likelihood parameters were estimated. A) Relative bias and 95% HPD width of the case count likelihood scaling factor *k*_*cc*_. The black, dashed line marks the 95% highest density interval of the prior distribution of the parameter. When case counts were excluded no case count likelihood related parameters were estimated. B) Relative bias and 95% HPD width of the case count likelihood dispersion *α*_*cc*_. C) Relative bias and 95% HPD width of the wastewater concentration likelihood scaling factor *k*_*ww*_. When wastewater concentrations were excluded no wastewater likelihood related parameters were estimated. D) Relative bias and 95% HPD width of the wastewater likelihood standard deviation *σ*_*ww*_.

**Supplementary Figure S9:**
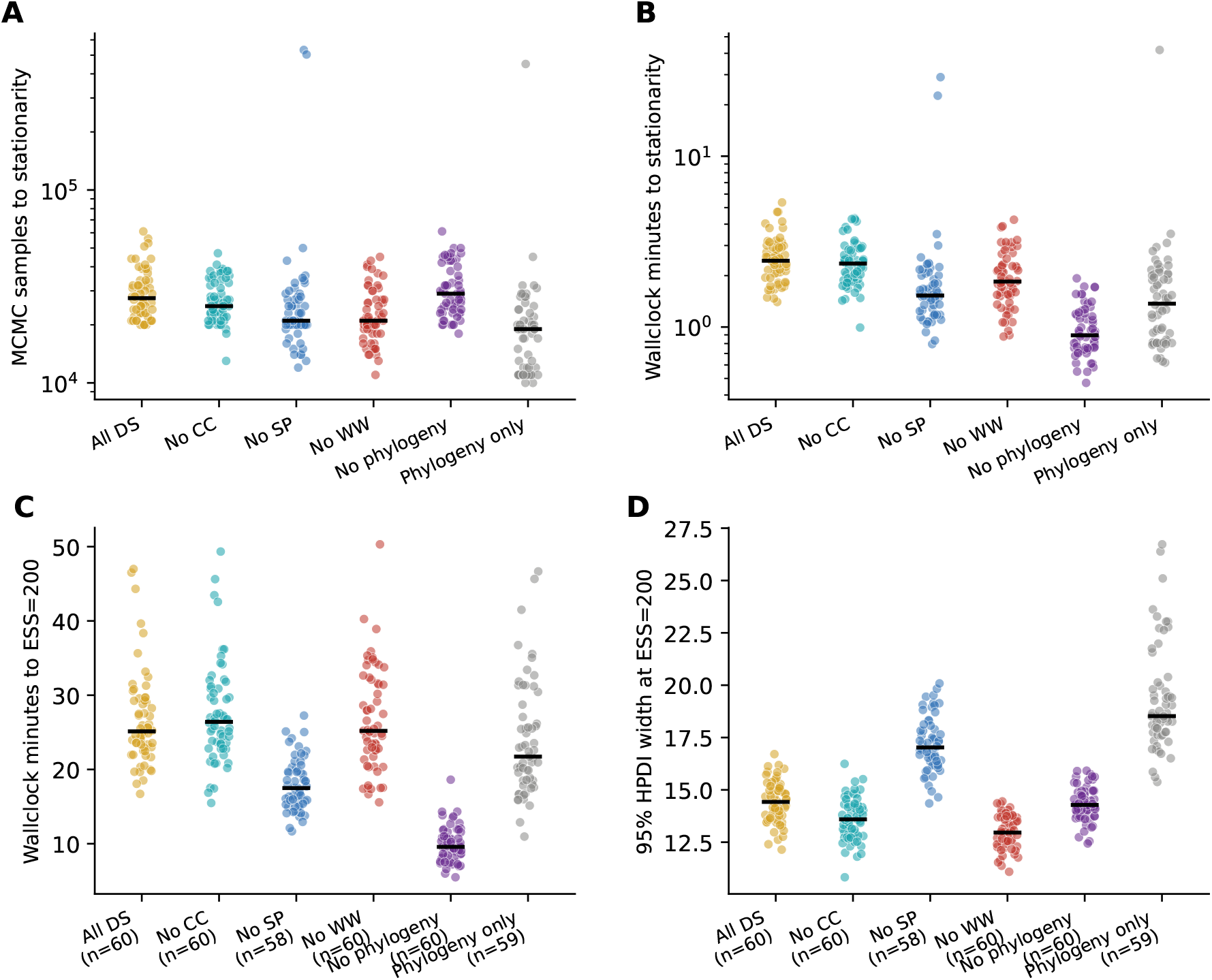
Computational cost of adding multiple data streams to MASCOT-DS for parameter inference assessed on simulated data. A) Number of MCMC samples necessary to reach stationarity of the posterior. Black solid lines indicate the median across 10 simulations x 3 seeds. Each dot shows the results for specific run. B) Wallclock time in minutes necessary to reach stationarity of the posterior. C) Wallclock time in minutes to reach the target effective sample size (ESS) of 200 for the posterior. The number of dots is given in brackets. Runs that did not reach 200 when they finished were not shown to help with visualisation. D) Width of the 95% HPD interval of the posterior MCMC chain when reaching ESS=200.

**Supplementary Figure S10:**
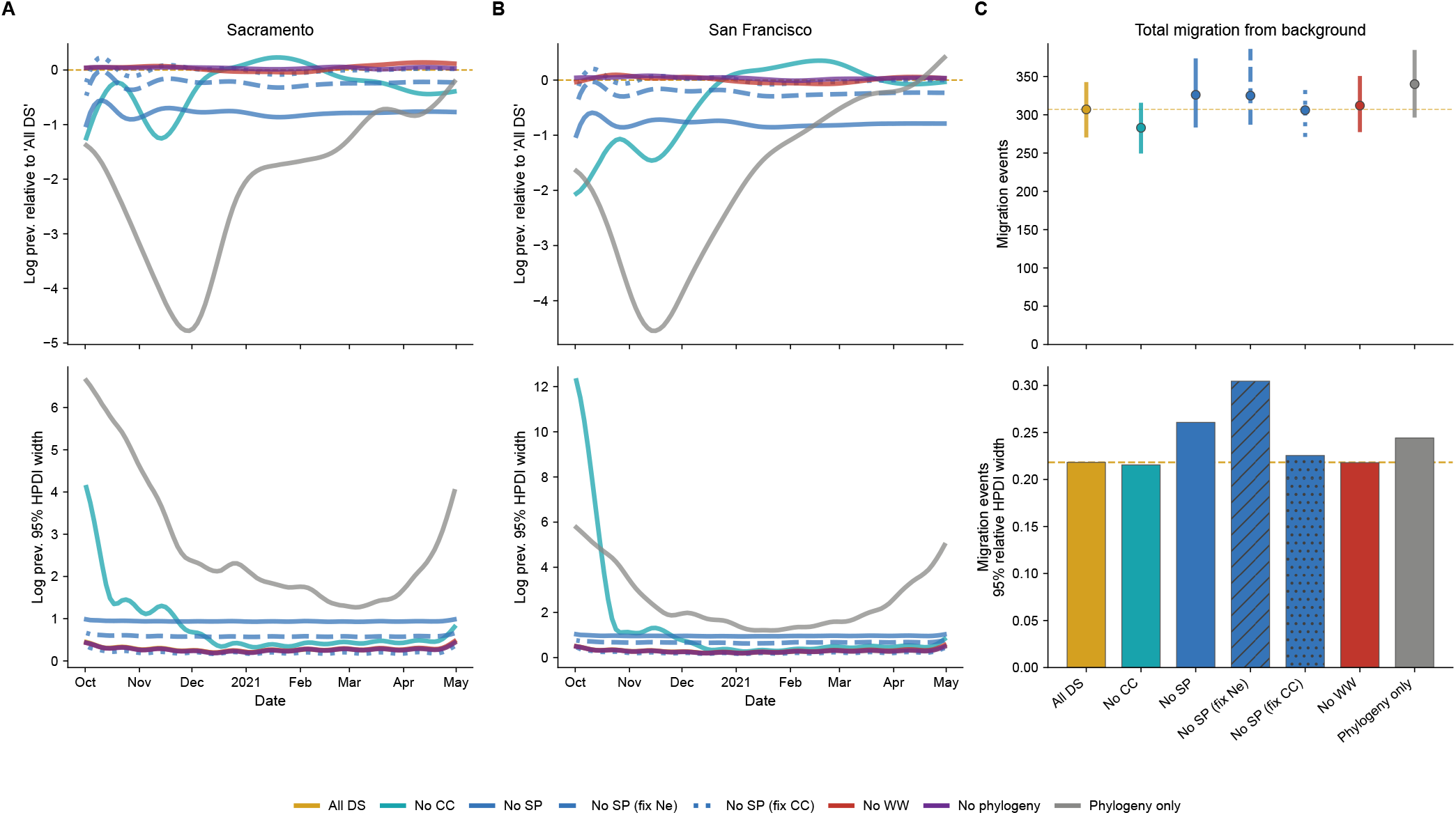
Value of data streams for reconstructing the SARS-CoV-2 winter 2020-21 wave in San Francisco and Sacramento. Each curve is one data stream input combination version as described in Figure 4. A,B) Bias relative to the “All DS” version and 95% HPD interval (HPDI) width of log prevalence trajectories for different data stream input combinations over time. C) Posterior distributions and 95% relative HPDI width (width divided by the posterior median) of the number of total migration events from the outside deme into the Bay Ara counties; dots are the median of the posterior distribution, whiskers span the 95% HDPI, dashed line indicates the median of the “All DS” version for reference.

**Supplementary Figure S11:**
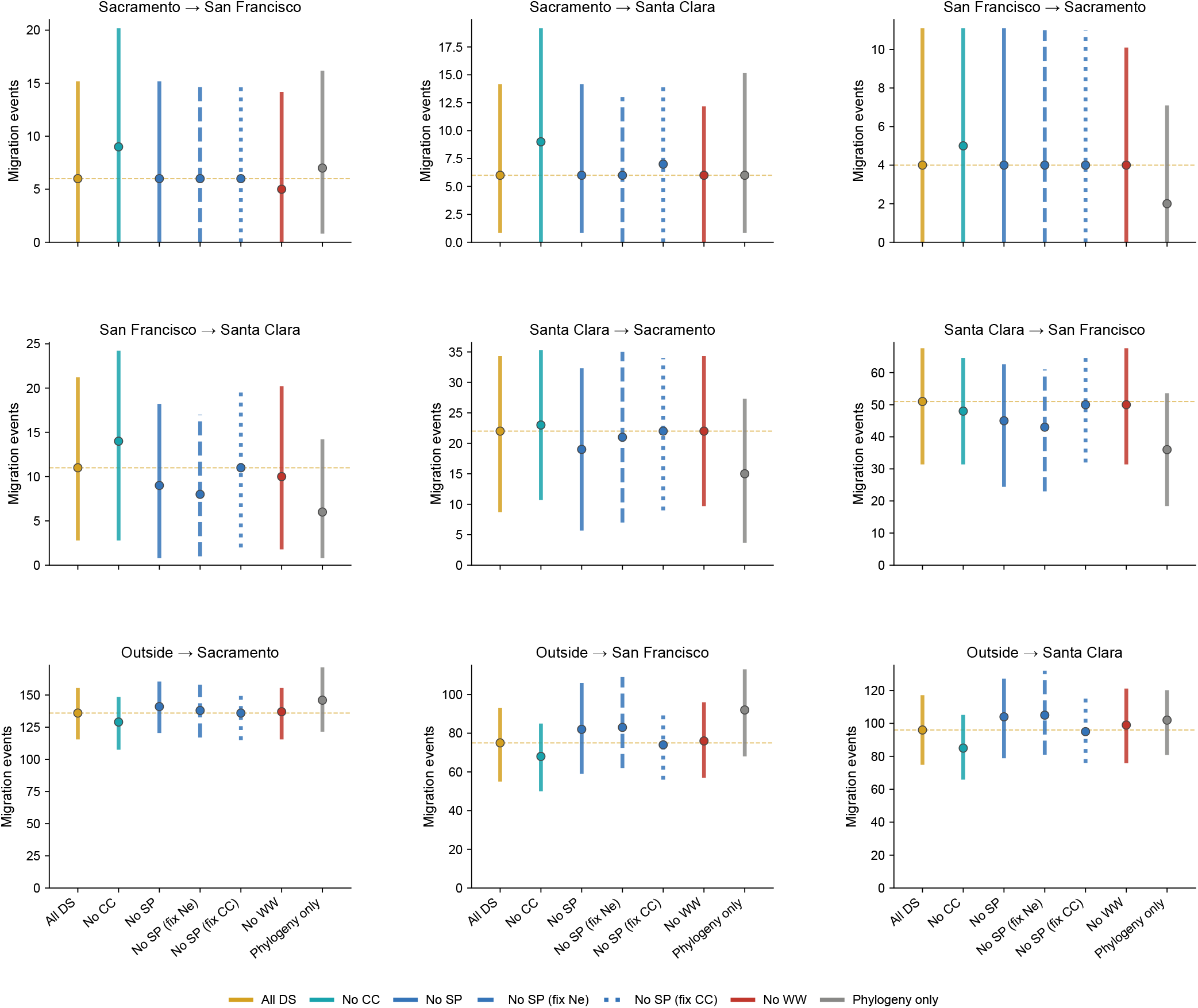
Inferred migration events between counties and with the background across inference variants, related to Figure 4. All estimated directional transitions among the three focal counties and the outside deme are shown; migration rates from counties into the background were not estimated. Variants are colored as in Figure 4. Posterior distributions of the number of migration events per direction and per variant, with source → destination counties indicated above each panel; whiskers indicate the 95% HPD, dots are the median of the posterior distribution.

**Supplementary Figure S12:**
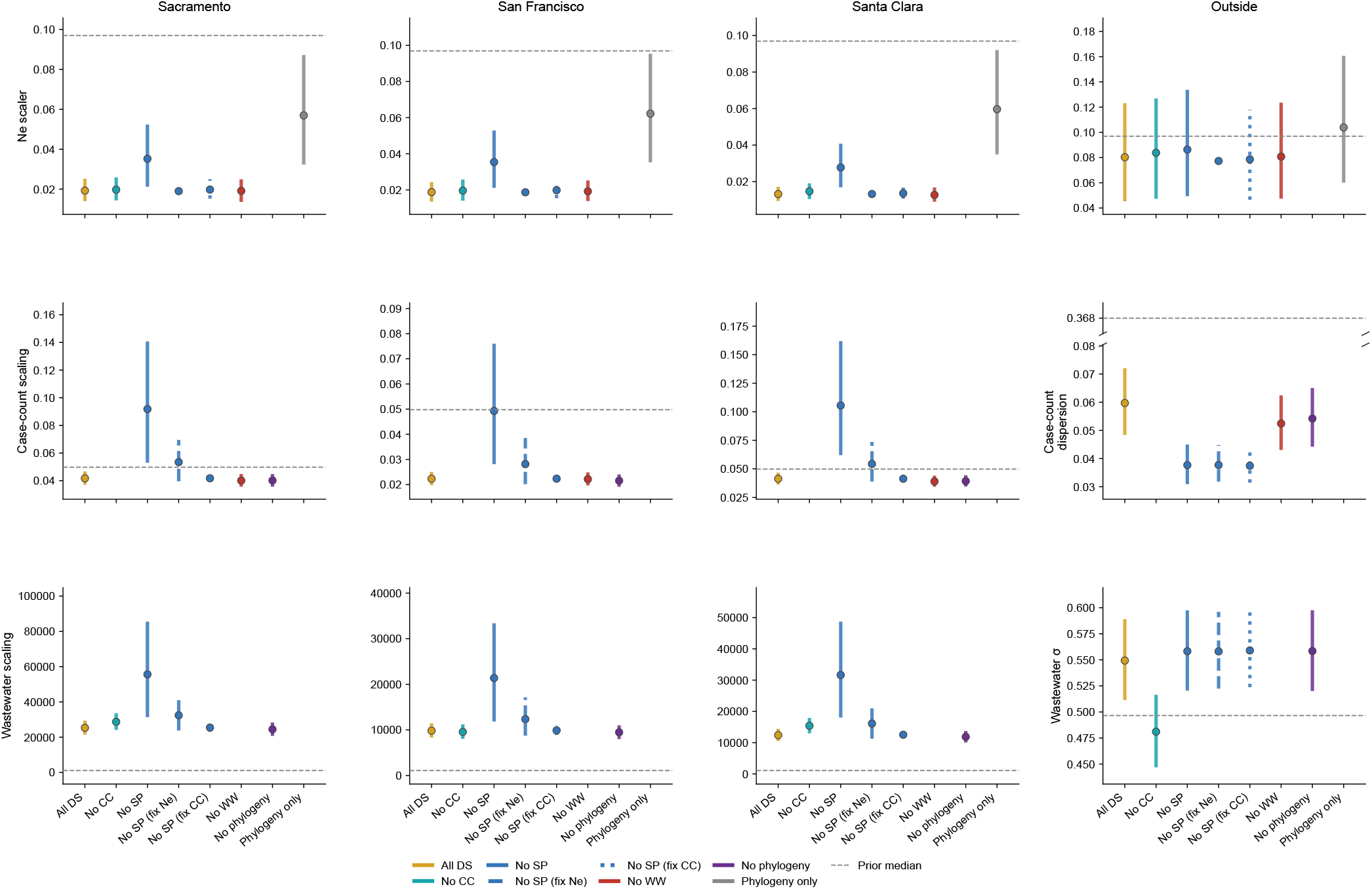
Posterior estimates of the data stream likelihood parameters across inference variants, related to Figure 4, variants that do not estimate a given parameter leave a gap. Grey, dashed horizontal lines mark the prior median of each parameter. First row contains estimates of effective population size (Ne) scaling factor per deme, including the outside deme. Middle row shows scaling factor and dispersion parameters of the case count likelihood. The dispersion panel uses a broken y-axis (//) so the much-larger prior mean and the tightly estimated posteriors are both visible. Bottom row shows scaling factor and standard deviation *σ* parameters of the wastewater likelihood. Dots are medians of the corresponding posterior distributions, whiskers indicate the 95%HPD interval.

**Supplementary Figure S13:**
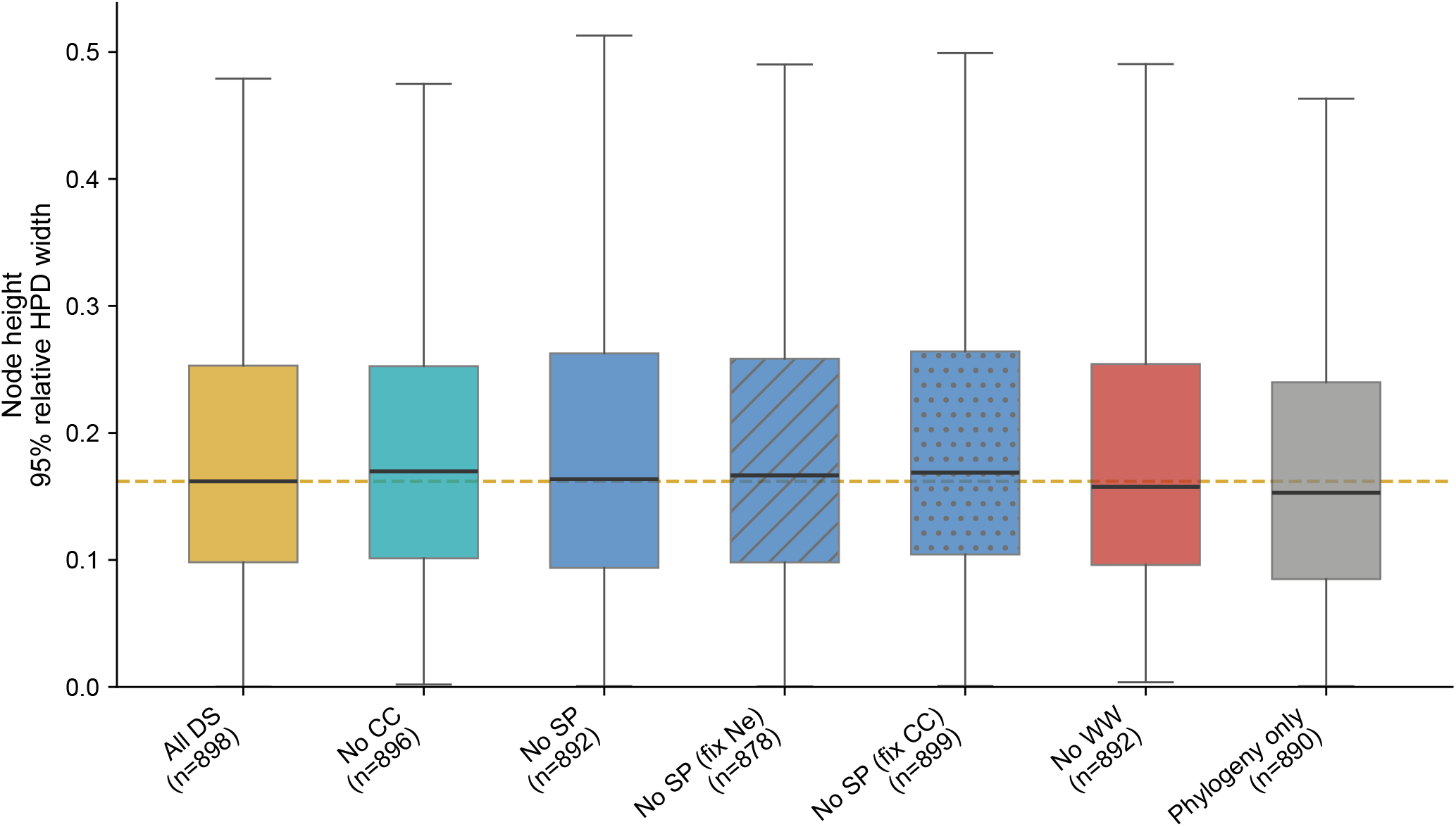
Uncertainty of node heights in the inferred phylogenetic tree of the SARS-CoV-2 winter 2020-21 wave compared across data stream input combination versions. For each input version, we extracted the relative 95% HPD intervals of node ages (95% HPD interval divided by the median) for all internal nodes as provided in the MCC tree annotation produced by treeannotator. Boxplots show the distribution of relative 95% HPD intervals of node ages (box = IQR, line = median, whiskers = 1.5*×*IQR, points = outliers). Yellow, dashed line shows the median of the “All DS” version. Treeannotator did not provide 95% HPD interval estimates for clades with very low posterior support, hence the number of nodes differs between input combination versions (given in brackets).

## Supplementary Methods

### Parameter inference using MCMC

#### MCMC proposals on prevalence knots

To make proposals to the prevalence knots more efficient, we implemented a suite of novel proposal distributions (“operators”). A joint up-down operator proposes a single Gaussian distributed *δ* in log space and applies it to all coupled parameters simultaneously: parameters already stored in log space receive an additive ± *δ* perturbation (a random walk), while parameters in real space are multiplied by exp(*±δ*) (a scaling move). A random walk operator that operates on a single index in a vector of parameters by adding a value *±δ*, sampled from a Bactrian distribution, to the parameter value of the selected index. A random walk operator that operates on a randomly selected range (“block”) of consecutive indices in a vector of parameters, by adding a value *δ*, sampled from a Bactrian distribution, to each parameter in the block, to aid efficient exploration of the prevalence trajectory parameter space.

#### Inference on simulated data

For each simulation, we built a MASCOT-DS XML configuration with this cleaned tree and the simulated data streams (case counts, wastewater viral concentrations, seroprevalence) as input. We pass the deme in which a leaf node was sampled to MASCOT-DS as type traits. The population sizes *N*_*i*_ and the becoming uninfectious rate *γ* were fixed to the values used in a given simulation.

##### Prevalence structured skyline parameterization

We parameterised the prevalence natural cubic spline with 11 knot times placed at equal intervals backwards in absolute time starting from the most recent sample to the root of the tree. We used a 1001-point regular grid to discretize the spline for faster likelihood computations. Because MASCOT-DS calculates transmission rates implicitly through the time derivative of the prevalence spline, downward jumps in prevalence can correspond to negative transmission rates. Log prevalence knot values were bounded between −15 and 15.

##### Priors

We applied the hard-constraint prior on the prevalence spline of each deme, rejecting MCMC proposals where transmission rates fall below zero. No further priors were used on the log prevalence knot values.

For data stream likelihood parameters, we used the sampling distributions as priors.

- Case count scaling factor *k*_*i,cc*_ ~ LogNormal(−3, 0.5)
- Case count dispersion parameter *α*_*cc*_ ~ LogNormal(−1, 0.5)
- Wastewater scaling factor *k*_*i,ww*_ ~ LogNormal(4.5, 0.5)
- Wastewater standard deviation *σ*_*ww*_ ~ LogNormal(−0.7, 0.3)

We used the sampling distribution of the cross-deme transmission rate *β*_*ij*_ as the prior on the forward migration rates, 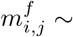 LogNormal(*µ* = 0.5, *σ* = 0.5)

##### MCMC inference configuration and proposal distributions

We performed MCMC inference for each simulation, with three different seeds and for 1,000,000 MCMC steps each, resulting in 3 *×* 100 = 300 total BEAST2 runs. To infer prevalence values, migration rates and data stream likelihood parameters we used the adaptable variance multivariate normal (AVMN) proposal distribution (“operator”) [67], as it is able to learn the correlation between parameters to make joint proposals for multiple parameters. We also used one up-down operator per deme to scale log prevalence up and case count and wastewater scaling factors down in joint proposals. We used Bactrian random walk operators [40] to propose updates to case count dispersion and wastewater concentration standard deviation parameters. For migration rates, we further utilized an adaptable operator sampler to choose between the AVMN proposal and a Bactrian scale proposal. Similarly, for prevalence knots of each deme, we utilized an adaptable operator sampler to choose between the AVMN proposal and a Bactrian random walk proposal. Additionally, we used the newly implemented Bactrian random walk block operator to propose updates to random contiguous blocks of prevalence knots.

##### Post-MCMC processing

For each simulation, we combined the results and trees of three runs using LogCombiner v2.7.7 with a 20% burn-in. We calculated the effective sample size (ESS) on the post-burn-in portion of each MCMC chain using a custom python script that calculated the autocorrelation of samples along the MCMC chain. We confirmed that ESS values were greater than 200 for the posterior, likelihoods and most parameters to ensure convergence.

#### Inference on SARS-CoV-2 data

We provided MASCOT-DS with the Epsilon variant multiple sequence alignment and the epidemiological data streams, case counts, wastewater viral concentrations and seroprevalence observations for each county. Additionally, we provided the population sizes *N*_*i*_ for each county (San Francisco: 892280, Sacramento: 1567975, Santa Clara: 1967585, obtained from the CDPH case count dataset). For the average infectious period we used seven days as a fixed parameter [68]. The outside deme was represented by the 70 background sequences across the US, but did not have any data streams or population size associated with it.

We used 13 knot points to parameterize the prevalence trajectory of each deme. Knot times were provided backwards in time, relative to the most recent pathogen sequence sampling date. For the Bay Area counties, we placed the first 11 knots starting from the most recent Epsilon sequence sampling date (04/30/2021) in regular intervals until 10/01/2020 in the past (start of time period of interest), then we added another knot two weeks earlier at 10/15/2020 and a last knot point at 04/30/2020 (one year before the most recent sample). These last two knot points allowed the prevalence estimates to explain coalescent events within counties outside of the time period of interest and to accumulate relevant infections if necessary to explain the data. We added two artificial “0” case count observations at these last two knot points to anchor the prevalence trajectory at low prevalence levels assuming no relevant Epsilon circulation. Since we expected the most recent common ancestor of all Epsilon sequences to have originated in the outside deme, we used a larger time span for the outside deme knot points to allow for reliable estimation of the phylogenetic tree root within a generous time range: we placed 12 evenly spaced knots from 04/30/2021 to 04/30/2020 and another knot at 04/30/2019, which is long before the emergence of SARS-CoV-2 [69] to provide a robust end point.

##### Priors

We applied a Normal(0, 1) smoothing prior on successive prevalence knot values of the outside deme to help regularise the prevalence trajectory of the outside deme. This prior was not needed for the Bay Area counties, since the prevalence trajectory was constrained by the additional data streams. We did not allow for migration from the counties into the outside deme. Thus only migration rates from the outside deme into the Bay Area and in between counties were estimated. We used a log-normal prior on the migration rates with a mean of 2 in real space and standard deviation of 1. For each deme, we estimated a scaling factor *k*_*i,Ne*_ since superspreading is prevalent in SARS-CoV-2 transmission [70]. We placed log-normal priors with a mean of 0.1 in real space and a standard deviation of 0.25 in log space on the scaling factors.

For each county, we estimated the scaling factor *k*_*i,cc*_ linking the prevalence trajectory to the expected case counts. We placed log-normal priors with a mean of −3 and a standard deviation of 0.5, both in log space, which was the same prior used in the simulation study. We used a single shared dispersion parameter *α*_*cc*_ across all counties, with a log-normal prior with a mean of −1 and a standard deviation of 0.5, both in log space, which was the same prior used in the simulation study.

For each county, we estimated the scaling factor *k*_*i,ww*_ linking the prevalence trajectory to the expected wastewater viral concentration. We placed log-normal priors on these scaling factors with a mean of 7 and a standard deviation of 1, both in log space, this was higher than the simulation study since we saw higher wastewater concentration values in the real data. We placed a single, shared log-normal prior on the standard deviation *σ*_*ww*_, with a mean of −0.7 and a standard deviation of 0.5, both in log space, which was the same prior used in the simulation study.

We fixed the clock rate to 0.001 mutations per site per year, to reduce the complexity of parameter inference. For the nucleotide substitution model, we used an HKY model with a log-normal prior on the transition-transversion ratio *κ*, with a mean of 1 and a standard deviation of 1.25, both in log space. Site-to-site rate heterogeneity was modelled with a discretised gamma distribution, with an exponential prior with a mean of 1 on the shape parameter. Nucleotide base frequencies were assigned a symmetric Dirichlet(4, 4, 4, 4) prior.

##### MCMC inference configuration and operators

We ran MCMC inference for 13 million steps, running the analysis in triplicate using different seeds. To infer the substitution-model parameters, we used an adaptable operator sampler [67] that alternates between the standard BEAST2 scale/exchange operators and the AVMN operator, allowing the sampler to learn which proposal mixes more efficiently for a given parameter. The same approach was used for the migration rates and the per-deme prevalence trajectories, where the AVMN operator learns the correlation structure between the migration rate estimates and between successive prevalence knot values within each deme. Additionally, we used the newly implemented Bactrian random walk block operator to propose updates to random contiguous blocks of prevalence knots for each deme. Since the prevalence trajectory of a deme and the case count and wastewater concentration scaling factors are correlated, we used the joint up-down operator that scales the prevalence knot values of a deme up while simultaneously scaling that deme’s case count and wastewater scaling factors down. The negative-binomial dispersion parameter of the case-count likelihood and the standard deviation of the wastewater observation noise were each proposed independently with a scale operator, without being coupled to any other parameter.

To propose new tree topologies and node heights, we used targeted tree operators that weight candidate moves by a consensus edge-support score computed across the sampled trees, focusing proposals on topological rearrangements and node-height changes that are more likely to be accepted compared to BEAST2 default tree operators [35].

##### Post-MCMC processing

For each simulation, we combined the results and trees of three runs using LogCombiner v2.7.7 with a 20% burn-in. We confirmed that ESS values were greater than 100 for the posterior, prior, tree likelihood, Mascot likelihood and most parameters to ensure convergence. We summarized the posterior samples of trees into a maximum clade credibility (MCC) tree while keeping the heights of the MCC tree using treeannotator.

### SIR simulation study using ReMASTER

We simulated an SIR outbreak involving two demes. The population size in both demes was set to *N*_*i*_ = 50,000 individuals. For each simulation, we randomly selected the outbreak start deme, thus the starting deme had *I*_*i*_(0) = 1 infected individual and *S*_*i*_(0) = *N*_*i*_ − 1 susceptible individuals at the start of the simulation. The becoming uninfectious rate *γ* was sampled from a uniform distribution *γ* ~ Uniform(50, 100)[yr^−1^] and the sampling proportion from Uniform(0.001, 0.002). We used time-varying reproductive numbers *R*(*t*) with three regimes: an initial high-transmission phase (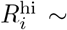 Uniform(2.0, 3.0)), a linear decrease phase, and a low-transmission phase (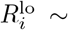 Uniform(0.2, 0.8)). Each regime occupies one third of a total period of *T*_*max*_ = 0.8 years. Within a simulation, *R*(*t*) was the same for both demes. Time-varying transmission rates within each deme *i* were calculated as *β*_*i*_(*t*) = *R*(*t*)*/γ*. To simulate cross-deme infection, we sampled the cross-deme transmission rate *β*_*ij*_ from a lognormal distribution *β*_*ij*_ ~ LogNormal(*µ* = 0.5, *σ*_*ww*_ = 0.5) for each cross-deme pair (*i, j*) with *i* ≠ *j*. We sampled one outbreak trajectory and phylogenetic tree per simulation under the structured SIR model in ReMASTER using *β*_*i*_(*t*) as the within-deme transmission rate, *β*_*ij*_ as the cross-deme transmission rate and *γψ* and *γ*(1 − *ψ*) as the recovery rates resulting in sampled and unsampled individuals, respectively. We ran each simulation using BEAST2 (v2.7.7) [40]. Simulations stopped when any of the following conditions was met: *T*_*max*_ was reached, the cumulative number of sampled individuals reached *n*_samples_ = 300 or no more infected individuals were present.

#### Computational cost of using different combinations of data streams in MASCOT-DS

We evaluated the computational cost of adding different combinations of data streams into parameter inference under the MASCOT-DS model. For this we defined two metrics of interest: the number of MCMC samples and wall-clock time required to reach stationarity, and the wall-clock time subsequently required to reach adequate mixing (effective sample size, ESS ≥ 200). We performed parameter inference on 10 SIR simulations, for each simulation varying the combination of input parameters: removing case counts (“no CC”), wastewater (“no WW”), seroprevalence (“no SP”) and the phylogeny (“no phylogeny”). In addition, we also tested the results when only providing SARS-CoV-2 Epsilon sequences as input (“Phylogeny only”). Each simulation and data stream input combination was run with three different seeds.

For each run and parameter, we defined the stationarity region as the 95% HPD interval across the last 50% of samples in the MCMC chain, an empirical estimate of its post burn-in posterior. Scanning forward from the first sample, we identified the earliest set of ten consecutive samples all falling within this region and took the corresponding last MCMC sample as the point of stationarity for that parameter. Because a chain is not stationary until its slowest-mixing parameter has stabilised, we defined run-level burn-in as the maximum stationarity point across all parameters of interest, and this burn-in was discarded before any downstream ESS or HPD interval calculation.

For each run and parameter, we evaluated ESS on a grid of MCMC-sample checkpoints along the post-burn-in chain, and recorded the wall-clock time, number of MCMC samples, and 95% HPD interval width at the first checkpoint where ESS reached 200. Wall-clock time was reconstructed from each run’s own average sampling rate (time per million samples, from the BEAST screen log). We excluded runs whose ESS never reached 200 within the sampled chain from this summary.

### Transmission rate derivation

MASCOT-DS calculates transmission rates *β* internally, these are not equivalent but related to transmission rates *β*^*SIR*^ used in an SIR model. We derive the definition of those transmission rates as the following.

In an SIR model the change in infected individuals *I*(*t*) at time *t* is given by the differential equation:

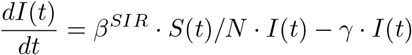

where *S*(*t*) is the number of susceptible individuals at time *t, N* is the total population size and *γ* is the becoming uninfectious rate.

In a multi-deme model, where we assume no movement of individuals between demes but cross-deme infections, 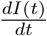 is given by the changes in local transmission and cross-deme transmission via migration rates *m*_*ij*_:

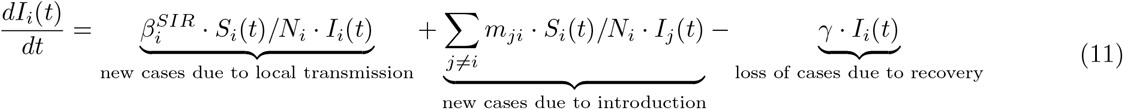

where *m*_*ji*_ is the migration rate from deme *j* to deme *i*.

MASCOT-DS derives the transmission rate *β* from the derivative of the prevalence spline (Eq. 1). Substituting 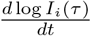 in Eq. 1 with Eq. 11 we get:

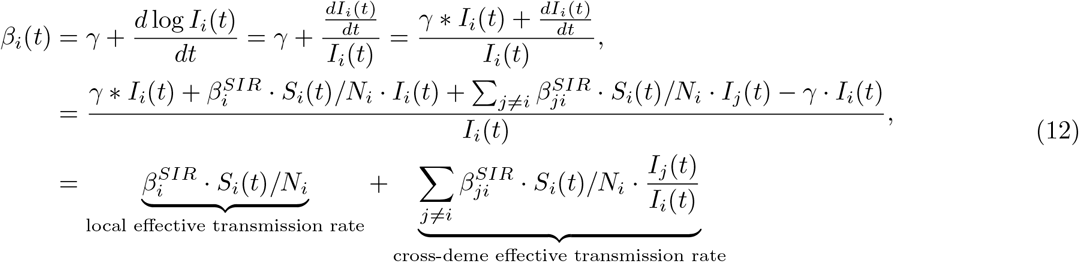

with 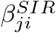 being the transmission rates from deme *j* to deme *i*.

Using migration rates as estimated by the MASCOT likelihood, which capture the rate a lineage migrates from deme *j* to deme *i*, we can define 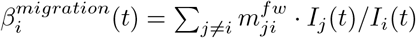 and we derive:

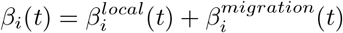

such that

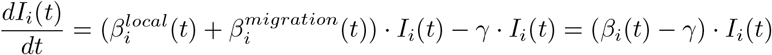

Thus, *β*_*i*_(*t*) can be understood as the effective transmission rate per infectious individual in deme *i*.

#### Caveat

In SIR terms:

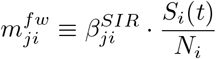

MASCOT-DS estimates 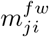 as a constant parameter, but 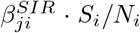 is time-varying as susceptibles deplete. The implicit assumption bridging these two is that 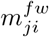 is a time-averaged or epidemic-phase-averaged effective importation rate, not a time-resolved quantity.

#### Relationship between *β* and effective population size

Based on [39] we can derive the relationship between *β* and the effective population size *Ne*_*i*_(*t*) as the following:

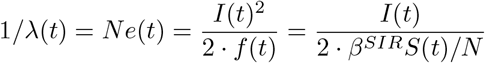

with *f* (*t*) = *β*^*SIR*^ *S*(*t*)*/N* · *I*(*t*) being the number of new cases produced locally. In the case of a one deme setting *Ne*(*t*) is exactly in MASCOT-DS:

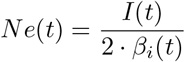

However, in settings with *n* demes, the coalescent rate between two lineages in deme *i* and *j* is given by:

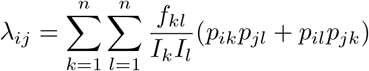

where *p*_*ik*_ is the probability of lineage *i* being in deme *k, f*_*kk*_ is the local birth rate 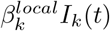 and *f*_*kl*_ is the cross-deme effective birth rate from *k* to *l* with 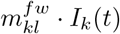.

If we are calculating effective population sizes in a multi-deme setting as

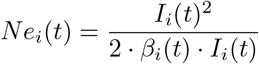

we underestimate *Ne*_*i*_(*t*) since 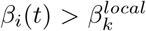 due to contributions from cross-deme transmission to *β*_*i*_(*t*) informed by the migration rates *m*_*ij*_.

If MASCOT is separately inferring migration rates *m*_*ij*_ from the tree, the same physical “cross-deme infection” event is contributing both to the inflated within-deme coalescent rate (via *β*_*i*_) and to the migration likelihood. The *Ne*_*i*_ posterior will be biased downward, and the migration-rate posterior might partly compensate.

MASCOT-DS has the option calculate the local transmission rate by using the total transmission rate and migration rate estimates

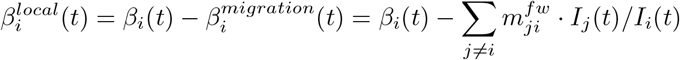

However, due to 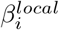 being required to be *>* 0 during MCMC proposals this places an effective bound on the migration rate posterior which might bias the estimates, especially when estimating fixed migration rates over the time period of interest. Additionally, MASCOT-DS cannot fit *I*_*i*_(*t*) = 0, so when there is truly no prevalence in a location MASCOT-DS will fit 0 *< I*_*i*_(*t*) *<* 1, which then numerically inflates 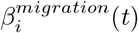 such that this in turn is another mechanism by which migration rate estimates can get biased to compensate the numerical issue in scenarios when *I*(*t*) *<* 1.

### Modeling bias of the infection hazard in the subpopulation tested in seroprevalence studies

We derive the seropositivity probability *p*_*i*_(*τ*) from the cumulative hazard Λ_*i*_(*τ*_*K*_, *τ*) experienced by a susceptible individual from the start of the outbreak *τ*_*K*_ to the observation time *τ* (Eq. 13).

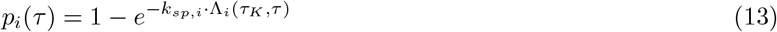

The scaling factor *k*_*sp,i*_ accounts for deme-specific differences in exposure risk of the tested population relative to the general population and can be fixed by the user or estimated through MCMC. To calculate Λ_*i*_(*τ*_*K*_, *τ*), we made use of the relationship of Λ_*i*_(*τ*_*K*_, *τ*) with cumulative incidence *C*_*i*_(*τ*_*K*_, *τ*) (Eq. 14).

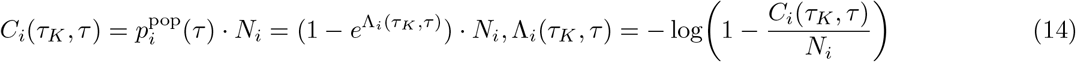

We compute the cumulative incidence *C*_*i*_(*τ*_*K*_, *τ*) from the prevalence spline grid by approximating integration of the incidence *β*_*i*_(*τ*)*I*_*i*_(*τ*) over time with the trapezoid rule on the prevalence evaluation grid (Eq. 15).

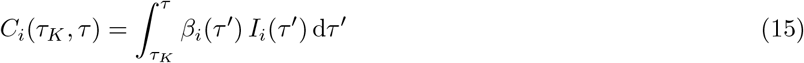

Substituting into Eq. 13 we can calculate the test-population specific probability of seropositivity *p*_*i*_(*τ*) (Eq. 16).

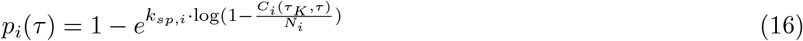

For numerical stability when 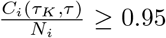, which can occur during MCMC exploration of the spline parameter space, log(1 − *C*_*i*_*/N*_*i*_) is replaced by its first-order Taylor extension at *C*_*i*_*/N*_*i*_ = 0.95.

