## Supplementary Appendix for "MASCOT-DS improves transmission dynamics inference by integrating multiple epidemiological data streams with phylodynamic inference"

All genome sequences and associated metadata supporting the findings of this study can be accessed through the persistent digital object identifier https://doi.org/10.55876/gis8.260725qm

In addition to the minted DOI, GISAID also communicates the aggregation of GISAID accession numbers (EPI_ISL_IDs) through the corresponding EPI_SET_260725qm identifier to facilitate both, the acknowledgment of all data contributors and the direct retrieval of the underlying data from GISAID used in this study.

### hCoV-19 Virus Data Summary

| GISAID Identifier | Digital Object Identifier | Number of individual viruses | Data Collection range | Number of countries/territories |
|---|---|---|---|---|
| EPI_SET_260725qm | https://doi.org/10.55876/gis8.260725qm | 999 | 2020-11-22 to 2021-04-30 | 1 |
